# Non-invasive measurement of intestinal barrier function in environmental enteropathy using transcutaneous fluorescence sensing

**DOI:** 10.1101/2025.11.05.25339488

**Authors:** Elena Monfort-Sanchez, Nilanjan Mandal, Mulima Mwiinga, Rose Banda, Jiacheng Xu, Monica N. Mweetwa, Tracy N. Phiri, Ian Chisenga, Jonathan Gan, Zixin Wang, Jingjing Qian, Arjun Agarwal, Ash Salam, María Gómez-Romero, Lynn Maslen, Elena Chekmeneva, Ara Darzi, Julian R. Marchesi, James Avery, Paul Kelly, Alex J. Thompson, the GI Tools Consortium

## Abstract

**Background:** Undernutrition represents a critical global health concern and is associated with a multifaceted breakdown in gut function – termed environmental enteropathy (EE) – which leads to increased intestinal permeability, inflammation and nutrient malabsorption. Current clinical approaches to assess intestinal permeability are costly, invasive, unreliable and/or difficult to perform in certain populations.

**Objectives:** We used transcutaneous fluorescence spectroscopy (TFS) – a novel method for non-invasive assessment of gut function – to investigate intestinal barrier function in EE.

**Design:** Volunteers were recruited in a Zambian community where EE is prevalent, and in the UK to undergo TFS measurements in a cross-sectional study. Data were compared between groups and were correlated with the lactulose:rhamnose (LR) test.

**Results:** TFS demonstrated significant differences in intestinal barrier function between UK and Zambian volunteers. Both peak fluorescence intensity (*p*=0.003) and area under fluorescence curves (*p*=0.02) were higher in Zambian than UK participants, suggesting increased permeation of TFS contrast agent. No differences were observed in time taken to reach peak, indicating no differences in factors affecting uptake rate (e.g. gastric emptying). Finally, fluorescence kinetics and regression analysis revealed strong correlations between TFS data and urinary recoveries of lactulose and rhamnose (Spearman’s *r* ≥ 0.78; *p* < 0.002).

**Conclusions:** TFS reveals population differences in permeability. It also allows simultaneous assessment of multiple elements of gut function (intestinal barrier integrity and gastric emptying) using a rapid, sample-free methodology. Combined with correlation to the LR test, this implies potential to advance studies of gut health and to improve clinical monitoring.

**KEY MESSAGES:** *What is already known on this topic:* Undernutrition represents a critical global health concern, accounting for nearly half of all deaths in children under the age of 5 and causing stunted physical and mental development in millions more. Importantly, undernutrition is hard to combat solely through provision of food as it is associated with a complex impairment in gut function known as environmental enteropathy (EE). A key facet of EE is so-called “leaky gut,” an effect where a degraded gut barrier allows passage of harmful compounds from the gut into the blood stream. Leaky gut has been widely studied in EE but understanding of the role of gut function in undernutrition is still incomplete due to the limitations of existing tests to measure gut leakiness (which typically involve measuring the recovery of sugar molecules in urine).

*What this study adds:* In this study, we used a new technology – transcutaneous fluorescence spectroscopy (TFS) – to study leaky gut in EE. TFS allows rapid, non-invasive assessment of gut leakiness by measuring the uptake of fluorescent molecules from the gut into the blood stream using a non-intrusive light sensor attached to the skin. Our results show that gut leakiness is higher in EE patients in Zambia than in healthy volunteers in the UK and that TFS results correlate with the lactulose:rhamnose (LR) test (an existing but much more cumbersome approach for measurement of gut leakiness).

*How this study might affect research, practice or policy:* Our results demonstrate that TFS can provide fast and non-invasive assessment of gut health in undernutrition settings. Due to its advantages over LR and other existing tests, this implies potential to allow more advanced study of EE and to improve clinical monitoring in the future.

## 1. INTRODUCTION

Undernutrition represents a crucial global health concern. The prevalence of undernutrition has continued to increase despite global efforts to combat hunger, and it remains a critical issue in certain regions, especially in sub-Saharan Africa and South Asia [1].

Importantly, numerous studies have demonstrated the surprising result that stunting (associated with chronic undernutrition) is not adequately corrected via nutritional supplementation alone [2–4]. In addition, undernutrition is associated with a complex, global breakdown in intestinal function known as environmental enteropathy (EE) [5–8]. EE comprises alterations in the mucosal structure of the gut (including villous blunting and increased crypt depth), increased gut (and systemic) inflammation, reduced absorptive capacity, an altered intestinal microbiome, and impaired intestinal barrier function [9, 10]. As such, EE is considered a central cause of the limited success of nutritional interventions in correcting growth faltering and has been associated with multiple poor socio-economic outcomes (e.g. low educational attainment, reduced income in adulthood, etc.) [2, 5, 11].

Impaired intestinal barrier function has been explored extensively in the context of EE and undernutrition. An aberrant gut barrier entails impaired tight junctions and epithelial breaks, which allow increased flux of large molecules from the gut into the blood stream via the paracellular and unrestricted pathways (**Fig. 1**). This leads to passage of bacterial peptides, bacterial cells, lipopolysaccharide (LPS) and other pathogen associated molecular patterns (PAMPs) into the bloodstream, triggering inflammatory responses that further exacerbate the underlying EE.

**Figure 1.**
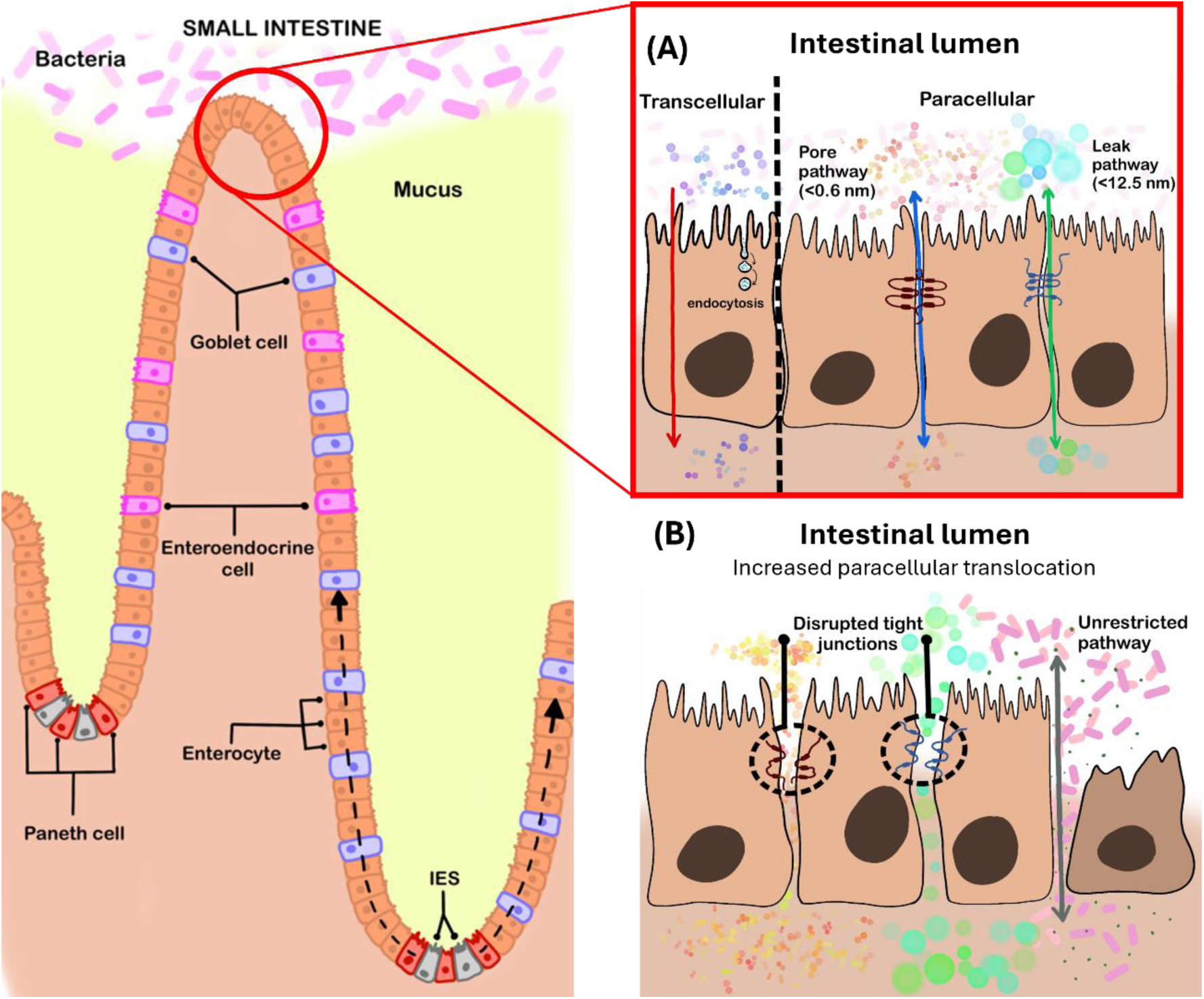
Intestinal barrier function in EE. Illustration of the cellular structure of the gut (left) and the transport of molecules (right) across **(A)** healthy and **(B)** compromised intestinal epithelial barriers (via transcellular, paracellular and unrestricted pathways). Intestinal epithelial stem (IES) cells control the renewal of the epithelial barrier. New epithelial cells migrate from the bottom of the crypt up the crypt–villus axis (dotted arrow) towards the villus tip. Secretory goblet cells and Paneth cells secrete mucus and antimicrobial peptides to protect the epithelial surface. In a healthy epithelial barrier **(A)**, the formation of a polarised cell monolayer results in the presence of an apical (upper) membrane in contact with the intestinal lumen and a basolateral (lower) membrane in contact with the neighbouring cells and connective tissues. The cells are linked together through tight junction complexes. Material can be transported across the gut barrier via transcellular or paracellular pathways. The transcellular pathway is an active transport mechanism mediated by transporters (i.e. endocytosis-exocytosis). The paracellular pathway is a passive transport mechanism where molecules pass through the gaps between cells (denoted as either the pore or leak pathway depending on molecular size). In a disrupted intestinal barrier **(B)**, integrity of the tight junctions is compromised increasing the inter-cellular spaces. Under more severe conditions, a third pathway (the unrestricted pathway) is generated, which allows large molecules to freely traverse the intestinal barrier in either direction.

A number of minimally- or non-invasive tests exist for clinical assessment of intestinal barrier function. These typically involve collection of urine or blood samples after oral ingestion of a test agent, and the most widely used methods are dual sugar assays such as the lactulose:mannitol (LM) and lactulose:rhamnose (LR) tests [12].

Dual sugar assays have been widely used in undernutrition studies (e.g. [13, 14]) due to the need for methods that can be deployed on large scales (e.g. for population and intervention-monitoring studies). Despite this, these existing assays suffer from a number of important limitations: they are cumbersome to perform (particularly in infants or patients with reduced urinary output); results exhibit considerable variability across studies; there is a lack of standardisation in how the tests are performed; and they require analysis of biological samples in advanced laboratories (leading to high costs, delays in reporting results, and risks of analytical batch effects) [13, 15]. As such, current protocols are not suitable for clinical assessment of intestinal barrier function at scale and/or in low-resource settings.

To address this issue, we have developed transcutaneous (‘through-the-skin’) fluorescence spectroscopy (TFS) for non-invasive monitoring of intestinal permeability and other gastro-intestinal (GI) functions [16–20]. This approach entails the oral administration of a clinically approved fluorescent contrast agent (fluorescein) and the use of a non-invasive fluorescence sensing probe (placed on the skin) to detect the permeation of that agent from the gut into the blood stream. We have previously demonstrated that TFS detects changes in both intestinal barrier function and gastric emptying rate [17, 18, 20]. Other human and animal studies have further demonstrated the potential of fluorescein and other fluorescent agents for assessment of intestinal barrier function in health and disease (e.g. [21–25]).

Importantly, TFS is a ‘sample-free’ method – it does not require collection of urine, blood or stool samples – and can thus produce clinical results within hours rather than weeks or months (as data can be analysed immediately without requiring laboratory analysis). The method also has potential to be low in cost and deployable on large scales due to the use of compact sensors, the use of fluorescein as contrast agent (with an established safety profile), and the lack of requirement for laboratory analysis. This indicates significant advantages for studies of EE, which is prevalent in low-resource settings where clinical and laboratory facilities often lack capabilities to perform existing tests.

Hence, in this study, we applied TFS (alongside the LR test) to a study of EE. Volunteers were recruited in both Lusaka, Zambia and London, UK to investigate differences in gut function between healthy and enteropathic groups. Our results demonstrate clear discrimination of the UK and Zambian cohorts as well as positive correlations between TFS and LR data. Together, this indicates the potential of TFS for large-scale, rapid and non-invasive assessment of gut function in low-resource settings, which will be useful in EE and many other health disorders characterised by a leaky gut.

## 2. METHODS

### 2.1. Clinical study recruitment

To study intestinal barrier function in EE, we developed and used two identical TFS sensors (see section 2.3). The first device (Sensor 1) was deployed in St. Mary’s Hospital (London, UK) to collect fluorescence data from 17 healthy volunteers (Group 1 – UK; participants UK1-17). The second device (Sensor 2) was deployed in St. Augustine Clinic (Lusaka, Zambia), where TFS experiments were performed in 34 volunteers (Group 2 – Zambia; participants Z1-34). Data from one UK participant (UK8) was excluded from analysis due to an aberrant fluorescence signal resulting from errors during data collection.

Measurements in the UK and Zambia were performed according to local clinical study protocols (which also provide sample size calculations; UK – [26], Zambia – [27]). Ethical approval was obtained through the UK Health Research Authority (HRA; IRAS Project ID – 242462; Research Ethics Committee (REC) reference – 18/LO/0714/AM04) and the University of Zambia Biomedical Research Ethics Committee (2291-2021, dated 20th December 2021) respectively.

All experiments were performed in accordance with Good Clinical Practice (GCP) guidelines and the World Medical Association’s Declaration of Helsinki. All volunteers gave informed consent prior to experiments, and all participants were aged 18 and above and had no previous reactions to fluorescein. Participants who were pregnant, breastfeeding, or had taken antibiotics within the last 4 weeks were excluded. Participant demographics – including age, sex, skin tone and body mass index (BMI) – are tabulated in **Tables S1** (UK) and **S2** (Zambia) (Supplementary Information).

### 2.2. Clinical data collection

All volunteers were seated/semi-recumbent for the duration of the experiments and all volunteers fasted overnight prior to the experiment. A TFS fibre probe was attached to the participant’s index finger. Fluorescence recordings were then started at the same time that the participant was asked to begin drinking a fluorescein solution containing 200 mg fluorescein dissolved in 100 ml water. Fluorescence signals were recorded for 180 minutes, with normalized fluorescence intensity values (fluorescence intensity divided by intensity of backscattered excitation light – see section 2.3 and [19]) measured every 20 s. This resulted in a fluorescence dataset containing 540 data points for each participant.

UK volunteers consumed a solution containing 200 mg fluorescein only. In Zambia, 20 participants consumed a solution containing 200 mg fluorescein in 100 ml water, and a separate subset of 14 participants consumed 200 mg fluorescein plus 5 g lactulose and 1 g rhamnose (in 100 ml water) [27]. This allowed comparison of TFS results against the LR test. Details of the LR testing protocol are presented in the Supplementary Methods (Supplementary Information).

### 2.3. Transcutaneous fluorescence sensors

Two compact, fibre-optic TFS sensors were used to measure transcutaneous fluorescence signals at the fingertip following oral ingestion of fluorescein (**Fig. 2**). The design and validation of the sensors (including optical/electrical safety considerations) have previously been described in detail [19]. Briefly, the sensors consist of: a 465 nm light emitting diode (LED) for excitation of fluorescence; two photodiodes for detection of fluorescence and backscatter signal intensities; optical filters to ensure high fidelity detection; a trifurcated fibre-optic probe to deliver and collect light from the skin; a spring-loaded, 3D-printed finger clip to ensure firm attachment of the fibre probe to the skin; electronic components (microcontroller and analogue-to-digital converter, ADC) for control of hardware; and a laptop PC running custom-written LabVIEW software for control of device, data acquisition and signal processing (**Fig. 2A-E**).

**Figure 2.**
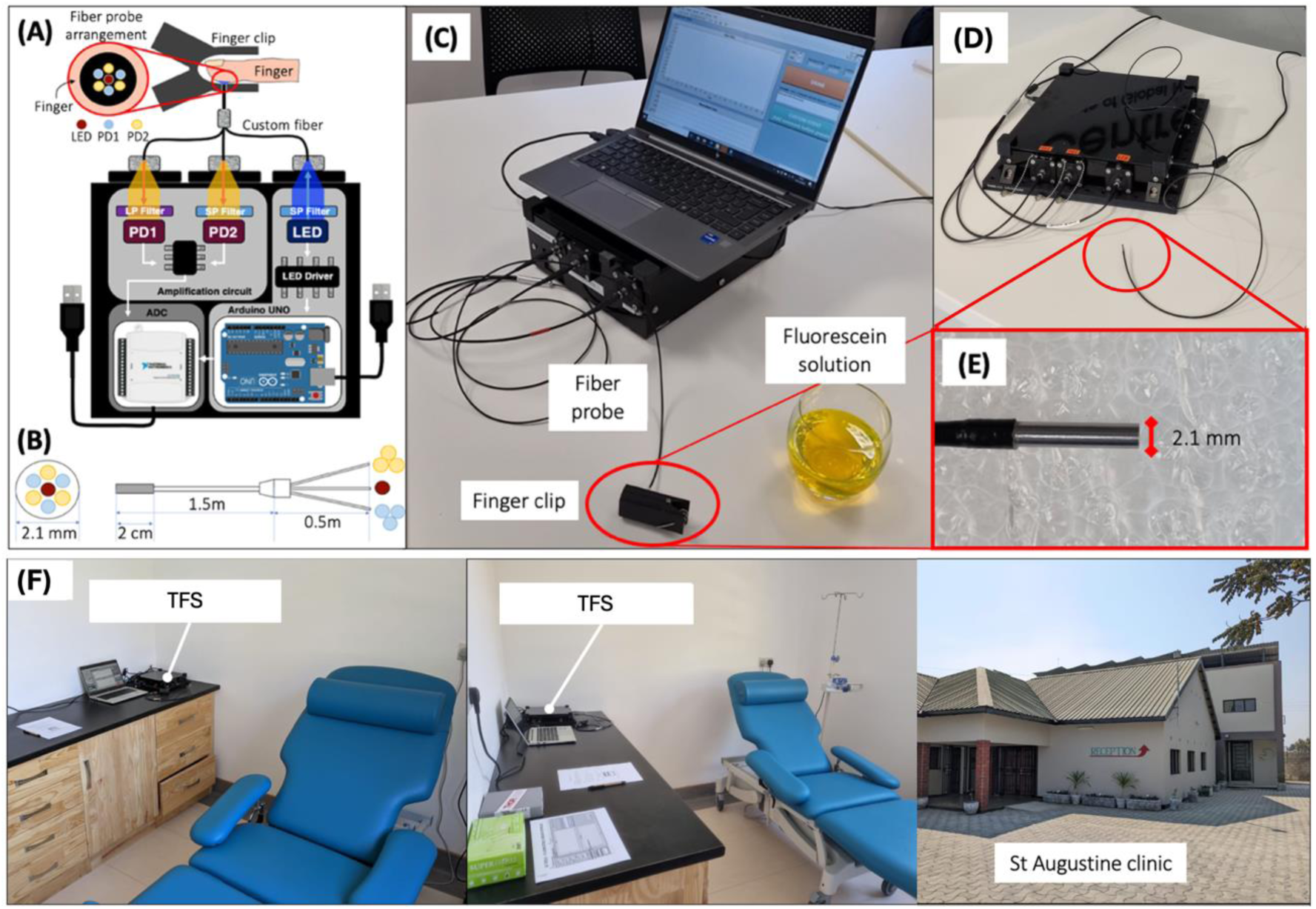
Compact fibre-optic fluorescence sensor for transcutaneous assessment of intestinal permeability. **(A)** Schematic diagram of compact TFS sensor. **(B)** Custom fibre probe diagram showing distal fibre arrangement (left) and full bundle (right). **(C)** Photograph of Sensor 1 (UK) showing: device housing; laptop running user interface for collection and pre-processing of data; fibre probe with 3D printed finger clip connected to fibre tip; fluorescein solution for oral ingestion. **(D)** Photograph of Sensor 2 (Zambia) showing device and fibre probe. **(E)** Close-up photograph of distal tip of optical fibre probe (which is identical for both sensors). **(F)** Sensor 2 deployed in clinical environment for TFS measurements (St. Augustine Clinic, Lusaka, Zambia).

The sensors record both the fluorescence and backscattered LED signal intensities at all time points. Those values are used to calculate a normalised fluorescence signal, thereby accounting for variations due to fluctuations in LED output, differences in skin tone, probe movement, and other factors that can affect signal levels [19]. The devices are compact, portable and user-friendly to enable straightforward use in clinical environments without the need for technical expertise. **Fig. 2F** shows Sensor 2 deployed in St. Augustine Clinic, Lusaka, demonstrating the small footprint and clinical compatibility.

To allow comparison of data collected with both sensors, each device was calibrated using a Fluorescence Standard (USFS-336-010, Labsphere Spectralon, Pro-Lite Technology Ltd., UK) that provided a known fluorescence output in response to 465 nm LED excitation (**Fig. S1&2**, **Table S3**, Supplementary Information). The calibration was further validated via *in vitro* fluorescence measurements on aqueous fluorescein solutions, which demonstrated good agreement between sensors following calibration (**Fig. S3**, Supplementary Information).

### 2.4. Data and statistical analysis

The collected data were analyzed to compare the transcutaneous fluorescence signals from Group 1 (UK) and Group 2 (Zambia) to investigate differences in intestinal barrier function. Correlations between TFS parameters and LR test results were also calculated. These analyses were first performed using single parameters extracted from TFS datasets (such as the peak intensity). Second, a physiologically based fluorescence kinetics (PBFK) model was developed to provide a full description of each TFS dataset. The fitted parameters from this model were used to compare Groups 1 and 2 and to explore correlations with LR parameters (via regression analysis). Full details of all data analysis protocols are provided in the Supplementary Methods (Supplementary Information).

### 2.5. Patient and public involvement

Experimental design was guided by public involvement activities in Zambia. As part of the protocol development, a community advisory board was assembled consisting of members of the public from local communities affected by EE. This community advisory board participated in a series of discussion groups to support study design. Following ethical approval, members of the advisory board also supported recruitment to the study by advertising within their local communities.

## 3. RESULTS

### 3.1. Fluorescence dynamics in UK and Zambian participants

All TFS datasets exhibited an increase in fluorescence intensity over time up to a peak point prior to a decay back towards zero, indicating permeation of fluorescein from the gut into the blood stream followed by elimination (**Fig. 3A**). Inspection of the mean fluorescence intensity curves (**Fig. 3B**) reveals clear differences between groups. In particular, Group 2 exhibits higher maximal fluorescence intensity than Group 1, suggesting a greater degree of fluorescein permeation.

**Figure 3.**
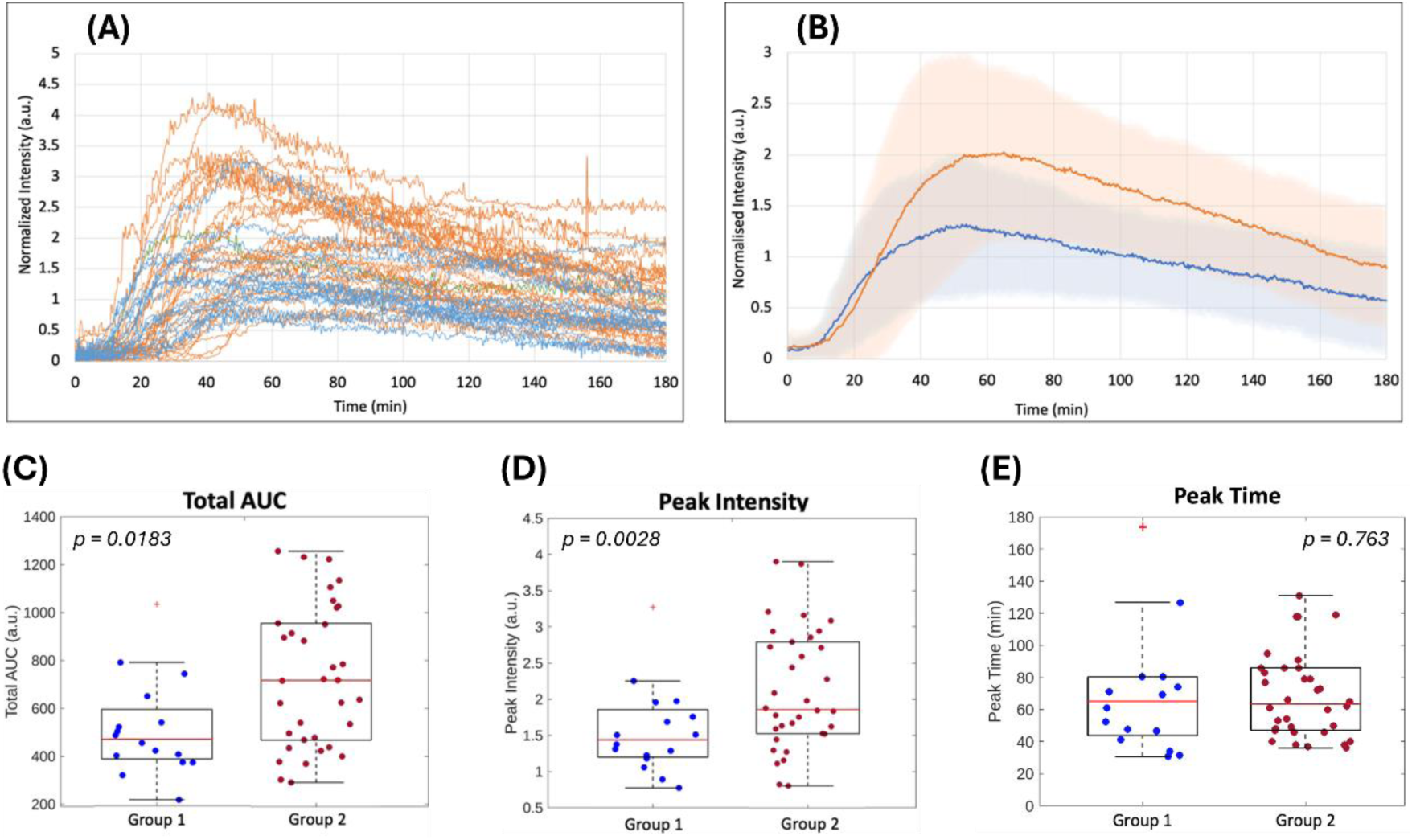
Analysis of fluorescence dynamics in UK (Group 1) and Zambian (Group 2) participants. **(A)** Normalized fluorescence vs. time curves for each participant for Group 1 (blue, n=16) and Group 2 (orange, n=34). **(B)** Mean normalized fluorescence vs. time curves for Group 1 (blue) and Group 2 (orange). Solid lines represent mean values, shaded regions represent error bars of ± one standard deviation. **(C-E)** Scatter-box plots displaying parameters extracted from the fluorescence vs. time curves: Total AUC **(C)**, Peak Intensity **(D)**, and Peak Time **(E)**. Box plots show median values (red lines), 25th and 75th percentiles (upper and lower bounds of black box), most extreme data points excluding outliers (whiskers), and outliers (red crosses). Blue and red circles represent individual values for Group 1 participants (blue) and Group 2 participants (red). Outliers were defined as points that fell below the 25th percentile or above the 75th percentile by more than 1.5 times the interquartile range. Importantly, outliers are shown for visualization purposes only, and all data points (including outliers) were included in all calculations and analysis. Inset p-values represent results of Wilcoxon Rank Sum tests comparing Groups 1 and 2.

To further explore differences between groups, a series of parameters – peak intensity, peak time, total AUC (area under curve), peak AUC (i.e. AUC from start of measurement up to the peak time), peak AUC / slope, and peak AUC * peak time – were extracted from each individual fluorescence curve (see Supplementary Methods, Supplementary Information). Both total AUC and peak intensity were significantly higher in Group 2 than in Group 1 (**Fig. 3C&D**; also see **Fig. S4&S5** and **Tables S4&S5**, Supplementary information), suggesting a greater degree of fluorescein permeation in Group 2.

Conversely, the time at which peak intensity was reached did not differ between groups (**Fig. 3E**). Despite a small apparent difference in peak time in the mean fluorescence curves (**Fig. 3B**), no difference was observed when calculating values across individual participants (**Fig. 3E**; see explanation of this effect in **Supplementary Note 1**, Supplementary Information). Similarly, the parameters peak AUC, peak AUC / slope, and peak AUC * peak time exhibited no statistically significant differences between groups (**Fig. S6**, Supplementary Information). This is likely explained by the similar peak times in each group (see **Supplementary Note 2**, Supplementary information).

Overall, these results suggest clear differences between UK and Zambian participants in terms of intestinal barrier function, but not in factors affecting rate of uptake (gastric emptying, gut motility, etc.).

### 3.2. Demographic analysis

Considering the significant difference in fluorescence intensity between UK and Zambian participants, we used the peak intensity parameter to further analyze data collected in the two populations, to investigate variability according to skin tone, sex, age, and BMI (**Fig. 4**; **Tables S6&S7**, Supplementary information; also see demographic information in **Tables S1&S2**, Supplementary Information).

**Figure 4.**
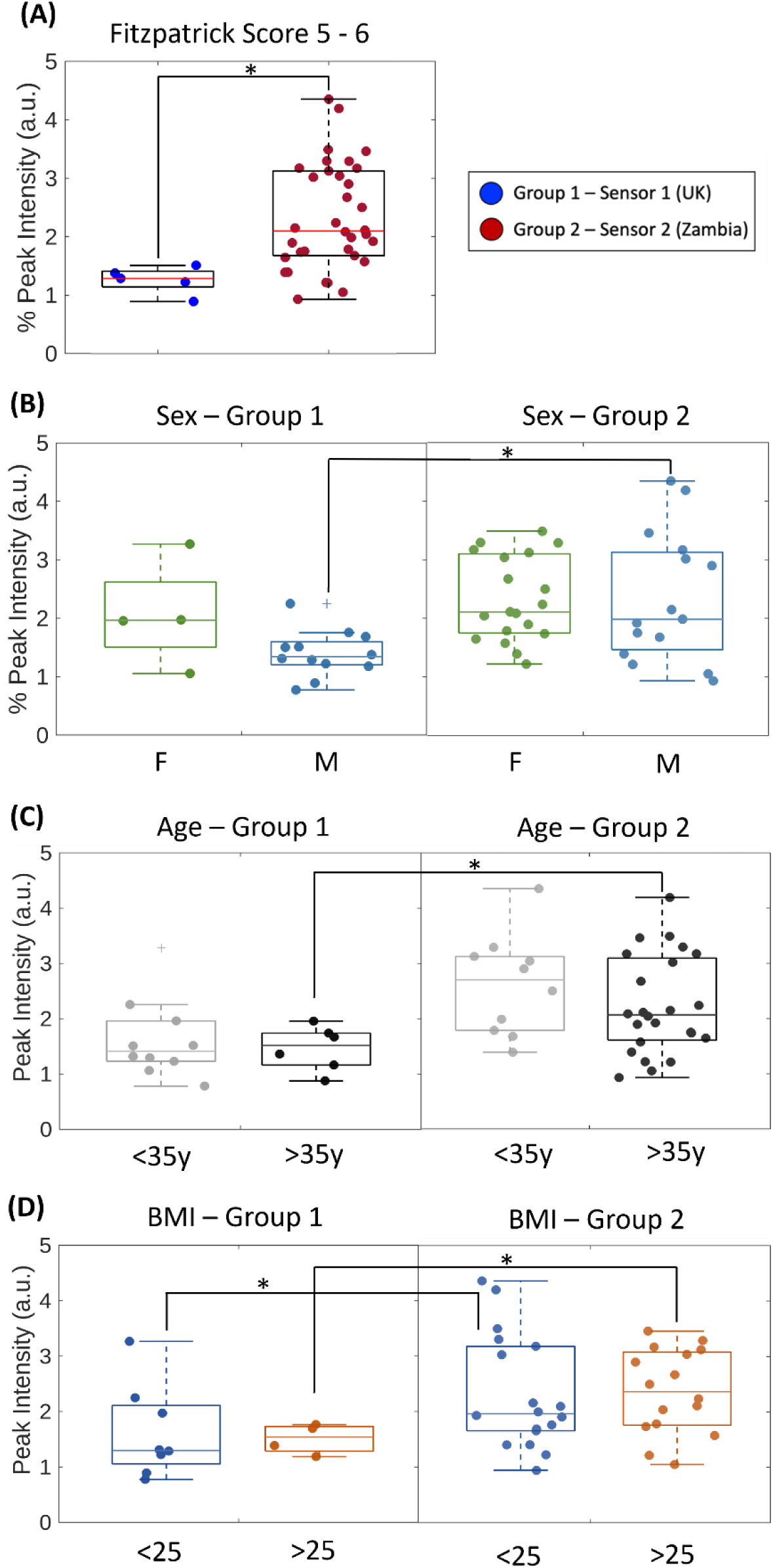
Demographic analysis. Scatter-box plots of the peak intensity values observed in each participant categorized according to skin tone, sex, age, and BMI. Box plots show median values (central horizontal lines), 25th and 75th percentiles (upper and lower bounds of boxes), most extreme data points excluding outliers (whiskers), and outliers (crosses). Coloured circles represent individual values. **(A)** Peak intensity values observed in participants with Fitzpatrick skin tone classes of 5-6 for Group 1 (blue) and Group 2 (red). Only Fitzpatrick classes 5-6 are shown, as Group 2 only contained participants in this class. **(B-D)** Peak intensity values grouped according to: **(B)** sex (F – female, M – male); **(C)** age (younger or older than 35 years old); and **(D)** BMI (lower or higher than 25). Asterisks indicate significance as assessed by Wilcoxon Rank Sum tests. * *p* < 0.05.

First, and importantly, when grouping participants according to skin tone (using the Fitzpatrick scale [28]), the significant difference in peak intensity between UK and Zambian participants was retained (**Fig. 4A**). This is consistent with previous data showing that transcutaneous fluorescence data was not reduced in the presence of darker skin tones [19].

When analyzing data according to sex, age and BMI, no significant differences were observed within Groups (i.e. within UK participants or within Zambian participants) between different categorizations (i.e. female vs. male, <35 years vs. >35 years, BMI<25 vs. BMI>25). Between groups (i.e. when comparing peak intensity values between Groups 1 and 2), however, significant differences were observed for males (but not females), for participants aged over 35 (but not under 35), and for participants in both BMI categories (>25 and <25) (**Fig. 4B-D**). This may indicate potential demographic differences in gut function, but it is important to note the low sample numbers used here (see further discussion in **Supplementary Note 3**, Supplementary Information).

### 3.3. Comparison against dual sugar permeability test

A subset of 14 volunteers in Group 2 took a LR permeability test at the same time as the transcutaneous fluorescence experiment. The mean urinary excretion of lactulose (%L) and rhamnose (%R) were 0.279% (range 0.023–1.329%) and 3.895% (range 0.253–12.635%) respectively, and the mean LRR was 0.082 (range 0.037–0.259) (**Table S5**, Supplementary Information). These %L and LRR values are higher than those typically reported in the literature for healthy subjects (e.g. [29]), as expected for an EE cohort.

We investigated correlations of all fluorescence parameters with LR results (**Table S8**, Supplementary Information). Linear regression suggested that peak intensity increased with increasing %L, %R and LRR (**Fig. 5**). Spearman’s Rank correlation analysis revealed a significant correlation between peak intensity and LRR (albeit at very close to the significance threshold; *r*=0.547, *p*=0.0459), but not for %L or %R (**Fig. 5**; **Table S8**, Supplementary Information). Total AUC also exhibited a statistically significant positive correlation with LRR (**Table S8**, Supplementary Information). While tentative due to the small number of measurements in this subset, these data may indicate that fluorescein translocation (as measured by peak fluorescence intensity or total AUC) is indicative of results from the LR test, a widely used marker of intestinal permeability.

**Figure 5.**
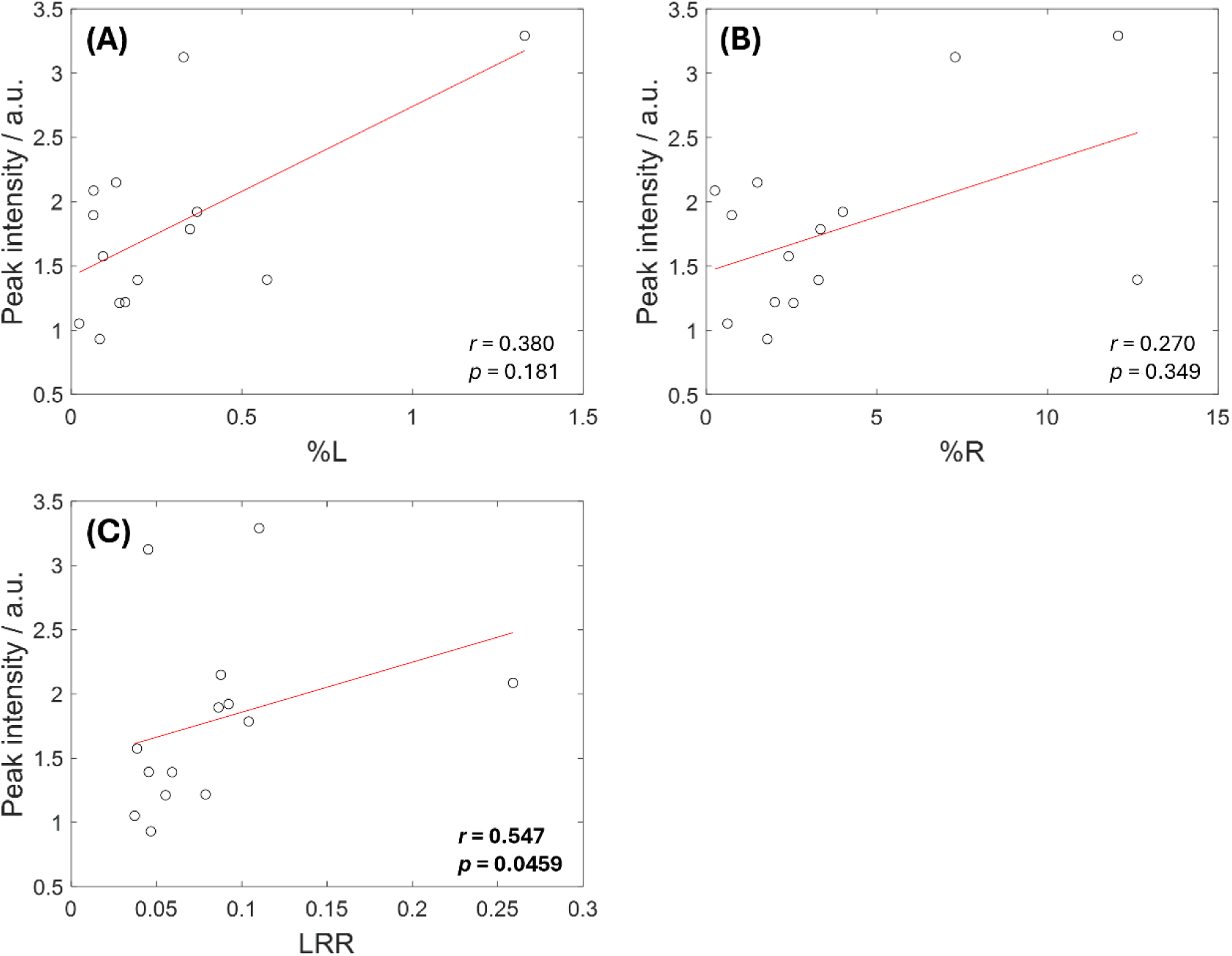
Correlations between peak intensity and LR parameters. **(A)** Peak intensity vs. %L. **(B)** Peak intensity vs. %R. **(C)** Peak intensity vs. LRR. Solid red lines indicate linear regression trendlines. Spearman’s Rank correlation coefficients (*r*) and corresponding *p* values for these trends are inset on graphs and presented in **Table S8** (Supplementary Information). Significant correlation (between peak intensity and LRR) is shown in bold.

### 3.4. Physiologically based fluorescence kinetics (PBFK) analysis

While the above results demonstrate the potential of TFS for non-invasive assessment of intestinal barrier function and reveal interesting trends between and within groups, all above analysis is based on interpretation of single parameters such as peak intensity. As such, this analysis may not provide a full assessment of the data as it discards a large proportion of the available information (i.e. by reducing datasets containing 540 data points into single parameters).

Therefore, we used kinetic/dynamic modelling to extract a set of parameters that provided a full description of the datasets. A PBFK model for orally ingested fluorescein was designed empirically (i.e. based on manual inspection of the fluorescence dynamics) incorporating two uptake and two elimination pathways – see details (including justification for use of a four-compartment model) in Supplementary Methods, Supplementary Information. This model provided considerable dimensionality reduction, reducing datasets from 540 data points to 9 fitted parameters. Crucially, however, the fitted parameters provided a full description of the observed data and, hence, allowed further analysis without loss of physiologically important information (i.e. PBFK fitting acted to remove noise while retaining key trends observed in the data).

The PBFK model exhibited good fits to all datasets, in cases with both one and two peaks in the fluorescence datasets (**Fig. 6A&B**; **Fig. S7&S8**, Supplementary Information). Residuals (difference between fitted curves and raw datasets) were typically randomly distributed around zero (**Fig. 6A&B**; **Fig. S7&S8**, Supplementary Information) and the mean (± standard deviation) *R^2^* goodness-of-fit parameter exceeded 0.96 in both participant groups (Group 1 – *R^2^* = 0.967 ± 0.021; Group 2 – *R^2^* = 0.971 ± 0.036). This demonstrates that our empirically designed model provided good fits to the data and, hence, a sound description of observed fluorescence dynamics.

**Figure 6.**
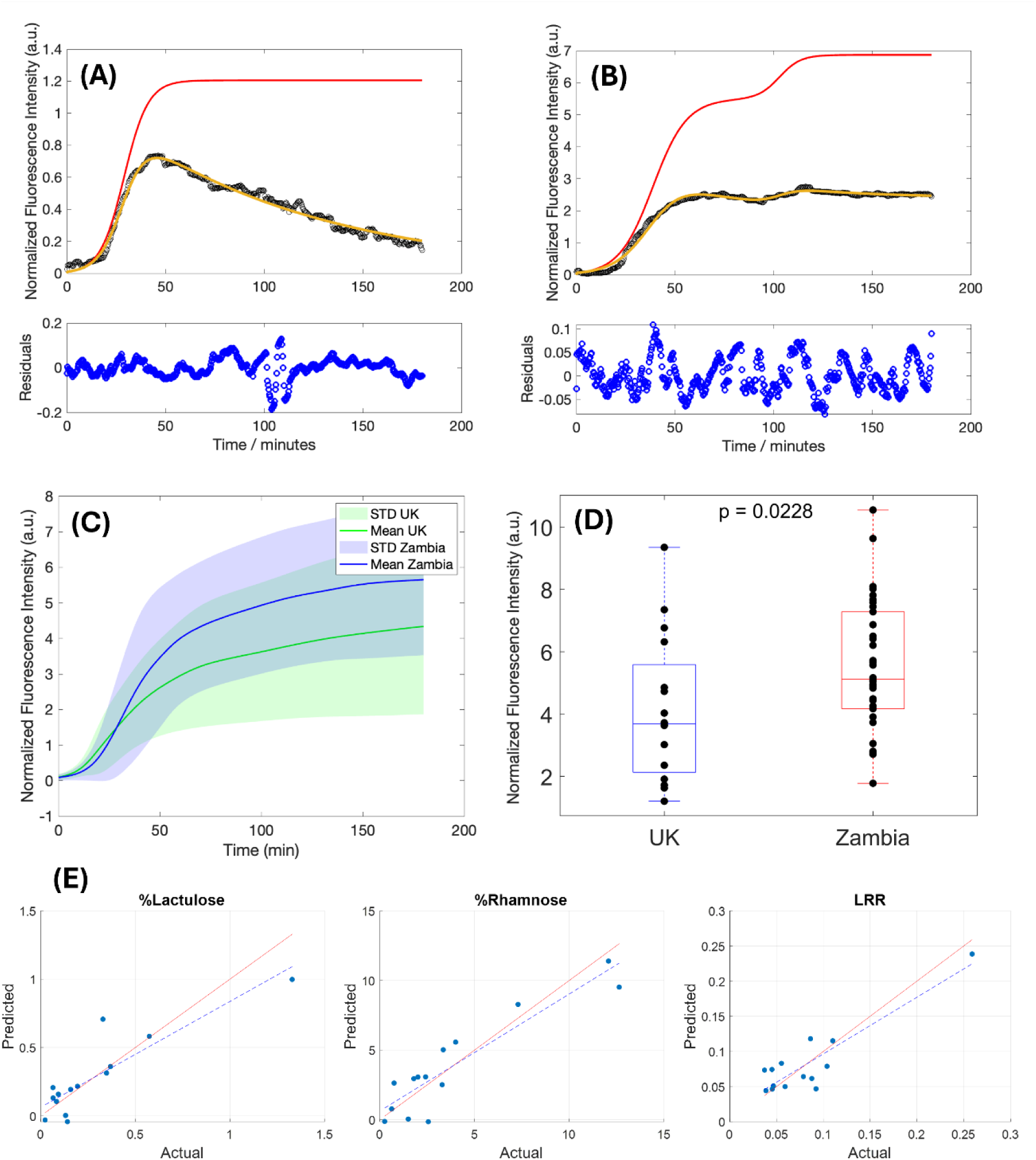
PBFK analysis reveals differences in total fluorescein uptake and correlation with the LR test. **(A, B)** Example PBFK model fits to datasets collected in a UK participant **(A)** and a Zambian participant **(B)**. Black circles represent raw data, yellow lines represent the fitted curves and red lines represent corrected “uptake-only” curves. Blue circles in subset plots represent residuals (differences between fit and raw data at all time points). Residuals appear randomly distributed around zero indicating good fits. **(C)** Average “uptake-only” fluorescence vs. time curves observed in UK and Zambia cohorts. Solid lines represent mean values, shaded regions represent error bars of ± one standard deviation (STD). **(D)** Scatter-box plot comparing the final fluorescence intensity values observed in uptake-only fluorescence curves (measured at 150 minutes). Box plots show median values (central horizontal lines), 25th and 75th percentiles (upper and lower bounds of boxes), and most extreme data points (whiskers). Black circles represent individual values. Inset p-value represents result of a Wilcoxon Rank Sum test comparing Groups 1 (UK) and 2 (Zambia). **(E)** Scatter plots comparing predicted and actual values for %L (left), %R (centre) and LRR (right). LR values were predicted using regression modelling with fitted fluorescence parameters as inputs. Dotted blue lines represent regression prediction trajectories. Red lines correspond to the y=x line (i.e. which would indicate perfect prediction).

Following fitting, the extracted PBFK parameters were used to calculate “uptake-only” fluorescence curves (see red curves in **Fig. 6A&B** and **Fig. S7&S8**, Supplementary Information). These uptake-only curves – which typically increased from zero towards a maximum/plateau point over time (**Fig. 6A&B** and **Fig. S7&S8**, Supplementary Information) – removed the effect of elimination and thus provided an estimate of the total amount of fluorescein absorbed. The mean uptake-only curves recorded in Groups 1 and 2 exhibited similar shapes but with higher fluorescence intensity observed in Group 2 (relative to Group 1) at all time points beyond approximately 30 minutes (**Fig. 6C**). The final fluorescence intensity in the uptake-only curves (measured at 150 minutes) was also found to be higher in Group 2 than in Group 1 (**Fig. 6D**). This further indicates a greater degree of fluorescein permeation (and, hence, impaired gut barrier function) in the Zambian cohort (relative to the UK cohort).

These results mirror those observed using the peak fluorescence intensity, which is attributed to the limited differences observed in peak time in this dataset – i.e. as the main differences between cohorts were observed in the intensity of the observed fluorescence signals and not in the time at which peaks were detected, the PBFK analysis revealed similar effects to the simpler peak intensity analysis. Importantly, however, this PBFK analysis further demonstrates the difference in gut barrier function between the two groups, but with a robust correction for any changes in gastric emptying, gut motility and rate of elimination.

### 3.5. PBFK-based prediction of LR parameters

Finally, in the subset of participants who took both TFS and LR tests, the fitted PBFK parameters and the final uptake-only fluorescence intensity were used in a regression analysis to explore prediction of LR parameters (i.e. %L, %R and LRR). Regression modelling (see Supplementary Methods, Supplementary Information) provided good fits to the LR data in all cases, with *R^2^* goodness-of-fit parameters of 0.77 or above (**Table 1**). Furthermore, the predicted %L and %R values extracted from the trained regression models were found to strongly correlate with the measured %L and %R values (**Fig. 6E**, **Table 1**; *r*≥0.780, *p*≤0.00158). A moderate but non-significant correlation was observed for LRR (**Fig. 6E**, **Table 1**; *r*=0.503, *p*=0.0694). For all LR parameters, mean absolute error (MAE) values were in the range of 20-40% of the mean observed LR values (**Table 1**).

**Table 1.** Linear regression modelling parameters. Goodness-of-fit (*R^2^*), Spearman’s Rank correlation coefficient (*r*), p-value for Spearman’s Rank correlation analysis (*p*), and mean absolute error (MAE) for linear regression fits of fitted PBFK parameters to %L, %R and LRR values.

| Parameter | $R^2$ | $r$ | $p$ | MAE |
| --- | --- | --- | --- | --- |
| %L | 0.774 | 0.780 | 0.00158 | 0.106 |
| %R | 0.841 | 0.793 | 0.00115 | 1.30 |
| LRR | 0.806 | 0.503 | 0.0694 | 0.0201 |

This demonstrates that when the full dynamics of the fluorescence curves are included in analysis, TFS datasets are strongly predictive of both %L and %R (with correlation to LRR also close to the significance threshold). This improves upon the results obtained when investigating correlations between the peak fluorescence intensity and LR parameters (**Fig. 5**), where only LRR was found to significantly correlate with the fluorescence parameter (with a p-value only fractionally below the *p*<0.05 significance threshold).

While the LR test and other dual sugar assays have limitations in assessment of intestinal barrier function, they nonetheless represent the most widely used tools for investigation of intestinal permeability in human disease studies. This is true in general but also particularly for studies of undernutrition and/or EE, where dual sugar assays have been deployed on large scales (e.g. [13, 14]) in part due to the challenges in obtaining other clinical data. Thus, the ability of TFS to provide a rapid, non-invasive measurement that is predictive of (and/or analogous to) an LR test indicates potential for significant benefit in global health settings by offering more reliable, more easily deployable, and much faster assessment of intestinal permeability.

## 4. DISCUSSION

We used non-invasive, transcutaneous fluorescence spectroscopy (TFS) to investigate intestinal barrier function in environmental enteropathy (EE). TFS revealed significant differences between healthy participants recruited in the UK and participants recruited in a community in Zambia previously shown to exhibit EE [30, 31]. In particular, the Zambian cohort were found to exhibit impaired intestinal barrier function relative to the UK cohort but no differences in gastric emptying or gut motility.

Both peak fluorescence intensity and total AUC were higher in Zambian than UK participants. This suggests a greater degree of permeation of fluorescein across the gut barrier in Zambian participants (relative to UK participants), implying impaired intestinal barrier function. This agrees with numerous previous studies reporting impaired gut barrier function in EE (e.g. [5, 6, 13, 32]).

Conversely, we observed no difference in fluorescence peak time between groups. This implies that there were limited or no differences between UK and Zambian participants in physiological factors that could affect the rate of uptake of fluorescein, such as gastric emptying and gut motility.

TFS parameters also correlated with the widely used LR test. First, peak fluorescence intensity (and total AUC) was found to significantly correlate with LRR (but not with %L or %R). Second, to further investigate correlations between TFS and LR data and differences between healthy and EE groups, we also performed kinetic modelling to provide a full description of fluorescence datasets (rather than reducing datasets to single parameters such as peak intensity). This analysis again revealed significantly higher fluorescein permeation in Zambian (relative to UK) participants. Furthermore, when coupled with regression analysis, fluorescence parameters extracted from kinetic modelling were found to be strongly predictive of both %L and %R.

Interestingly, these observations may be partially explained by a correlation between the recoveries of lactulose and rhamnose (**Fig. S9**, Supplementary Information). We observed a strong correlation between %L and %R (**Fig. S9A**, Supplementary Information), suggesting that both lactulose and rhamnose permeate the gut barrier to greater degrees as the epithelial barrier degrades. This is counter to the proposed mechanism of the LR test: rhamnose is chosen as a small molecule that is expected to permeate the healthy gut barrier. Thus, %R is expected to remain constant as barrier integrity degrades (or even decrease indicating reduced absorption). In this way, it acts as a normalizing agent for %L, allowing the LRR to provide a correction for variability in urinary excretion levels due to differing gut motility, gastric emptying or elimination rates between participants. However, in this case we observed that %R increases with increasing %L (**Fig. S9A**, Supplementary Information). This results in LRR being approximately constant with respect to both %L and %R (**Fig. S9B&C**, Supplementary Information). This is a crucial observation as it suggests that LRR is not a good marker of changes in intestinal permeability (i.e. as it in fact appears to mask changes in urinary lactulose recovery). This aligns with work by Ordiz et al., who reported similar findings using the LM test – i.e. that use of lactulose alone is equivalent or superior to use of a ratiometric measurement [33].

Importantly, these results may explain why our kinetic modelling and regression analysis revealed significant correlations for %L and %R but not for LRR. Our data suggests that TFS fluorescence intensity, %L and %R all increase with respect to increasing gut barrier degradation, but that LRR remains approximately constant (i.e. LRR does not correlate with %L – see **Fig. S9B**, Supplementary Information; also note tight clustering of all but one LRR value in **Fig. 6E**). Thus, TFS can be expected to be predictive of %L (and %R) but not LRR, as we observed here. Interestingly, TFS-based prediction of %L may be further explained by the similar molecular weights of lactulose and fluorescein (342 and 332 g/mol respectively), which may suggest that the two molecules would permeate the gut barrier to similar degrees under given conditions.

Finally, the main limitations of this study include the limited sample size (34 EE participants, 17 healthy participants) and the small number of participants in the subset who took both TFS and LR tests.

Overall, these results suggest exciting potential for TFS to improve upon dual sugar assays for assessment of intestinal barrier function by providing rapid, non-invasive and more reliable measurements. In addition, as TFS measurements are made with a high time resolution (i.e. one measurement every 20 seconds in the data reported here), they also allow for extraction of a higher level of physiological information (e.g. allowing simultaneous assessment of intestinal permeability and gastric emptying). Together, this suggests potential for significant impact in global health settings, particularly for study of EE, where dual sugar tests are widely used but suffer from numerous limitations. Incorporating TFS into studies of EE (and other gut diseases) may allow for both larger scale deployment and more advanced study, thereby enhancing understanding of this important and widespread condition.

## Acknowledgements

This work was supported by the UK Medical Research Council (MRC) through the GI Tools Project (grant reference: MR/V012452/1). The authors acknowledge both financial support from the GI Tools Project as well as intellectual input from all members of the GI Tools Consortium, which helped to guide and support protocol development, experimental design, data collection and data analysis. All members of the GI Tools Consortium are listed below.

## GI Tools Consortium

Beatrice Amadi, Rosemary Banda, Ellen Besa, Claire Bourke, Mutsa Bwakura-Dangarembizi, Ian Chisenga, Christine Edwards, Gary Frost, Isabel Garcia Perez, Mahek Jain, Leolin Katsidzira, Lydia Kazhila, Paul Kelly, Mirriam Kunaka, Kathryn Maitland, Nilanjan Mandal, Julian R. Marchesi, Callum Melvin, Elena Monfort-Sanchez, Douglas Morrison, Monica N. Mweetwa, Mulima Mwiinga, Perpetual Ngalande, Tracy N. Phiri, Joram M. Posma, Ruari Robertson, Jose Ivan Serrano Contreras, Aaron Konzani Tembo, Alex J. Thompson, James W. Weatherill.

LC-MS/MS method development work was supported by the Medical Research Council UK Consortium for MetAbolic Phenotyping (MAP UK) [grant reference: MR/S010483/1]. Infrastructure support was provided by the National Institute for Health Research (NIHR) Imperial Biomedical Research Centre (BRC). The views expressed in this publication are those of the authors and not necessarily those of the NHS, the National Institute for Health Research or the Department of Health. J.A. acknowledges an Imperial College Research Fellowship.

## Author Contributions

PK and AJT conceived the study. AJT, PK, EMS, MM, RB and JA designed the experiments. PK, MM, RB, AJT, JG and EMS prepared ethics application documents. EMS and JA developed the TFS data collection system. EMS, MM, RB, PK, NM, JG, JQ and AA performed experiments. EMS, JX, ZW, NM, JA and AJT performed data and statistical analysis. EC, AS, MGR and LM developed the LC-MS/MS method for urinary L:R analysis and performed all LC-MS/MS measurements. JA, AD and AJT contributed PhD student supervision. EMS, NM, JG, JA and AJT provided MRes student supervision. PK, AJT and AD secured funding. EMS and AJT wrote the manuscript. All authors edited and approved the manuscript.

## Data Availability

Data underlying the results presented in this paper are not publicly available at this time but may be obtained from the corresponding author upon reasonable request.

