## Supplementary Information for "Non-invasive measurement of intestinal barrier function in environmental enteropathy using transcutaneous fluorescence sensing"

**This PDF file includes:**

1. Supplementary Figures S1-S9
2. Supplementary Tables S1-S9
3. Supplementary Methods
4. Supplementary Notes 1-3
5. References

### 1. Supplementary Figures

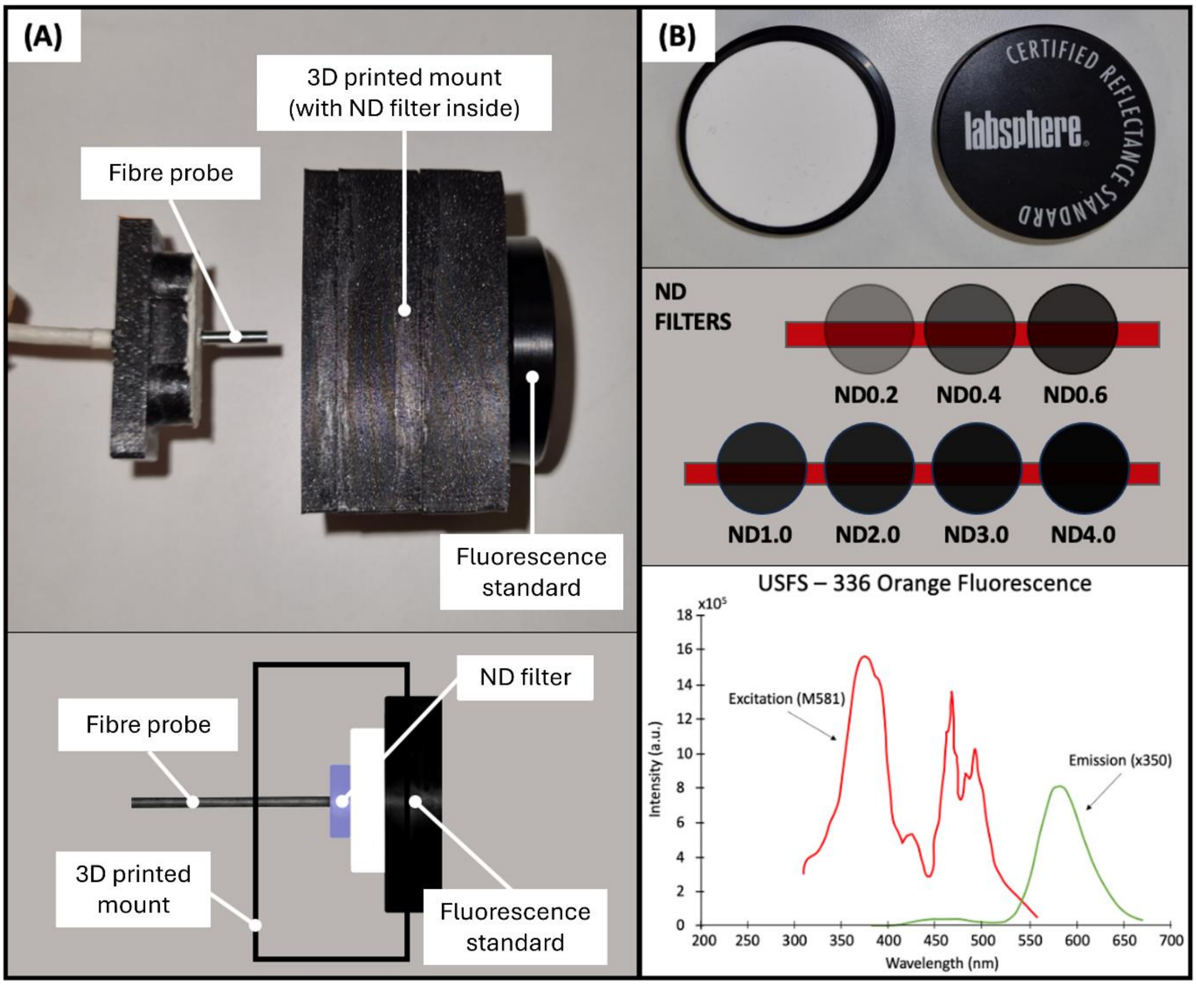


**Figure S1.** Sensor calibration experiments. **(A)** Experimental setup for calibration experiments: photograph (top); schematic diagram (bottom). Fibre probe (with the finger clip removed) is placed inside a 3D printed mount. The 3D printed mount prevents background light affecting the measurements. The fibre probe tip is positioned perpendicular to the fluorescence standard with a neutral density (ND) filter positioned between the probe tip and the fluorescence standard to attenuate light. Measurements were made using a series of ND filters to determine an attenuation level that provided signal levels within the sensitive range of the detection photodiodes (i.e. to obtain signal levels similar to those obtained *in vivo*). **(B)** Photograph of fluorescence standard (top); Illustration of ND filters used in calibration measurements (middle); graph showing excitation and emission spectral profiles (as specified by manufacturer) for the fluorescence standard used for calibration (bottom). In middle image, an ND filter value of "NDX" corresponds to an attenuation level of $1/10^X$ (i.e. it represents a filter with an optical density of X).

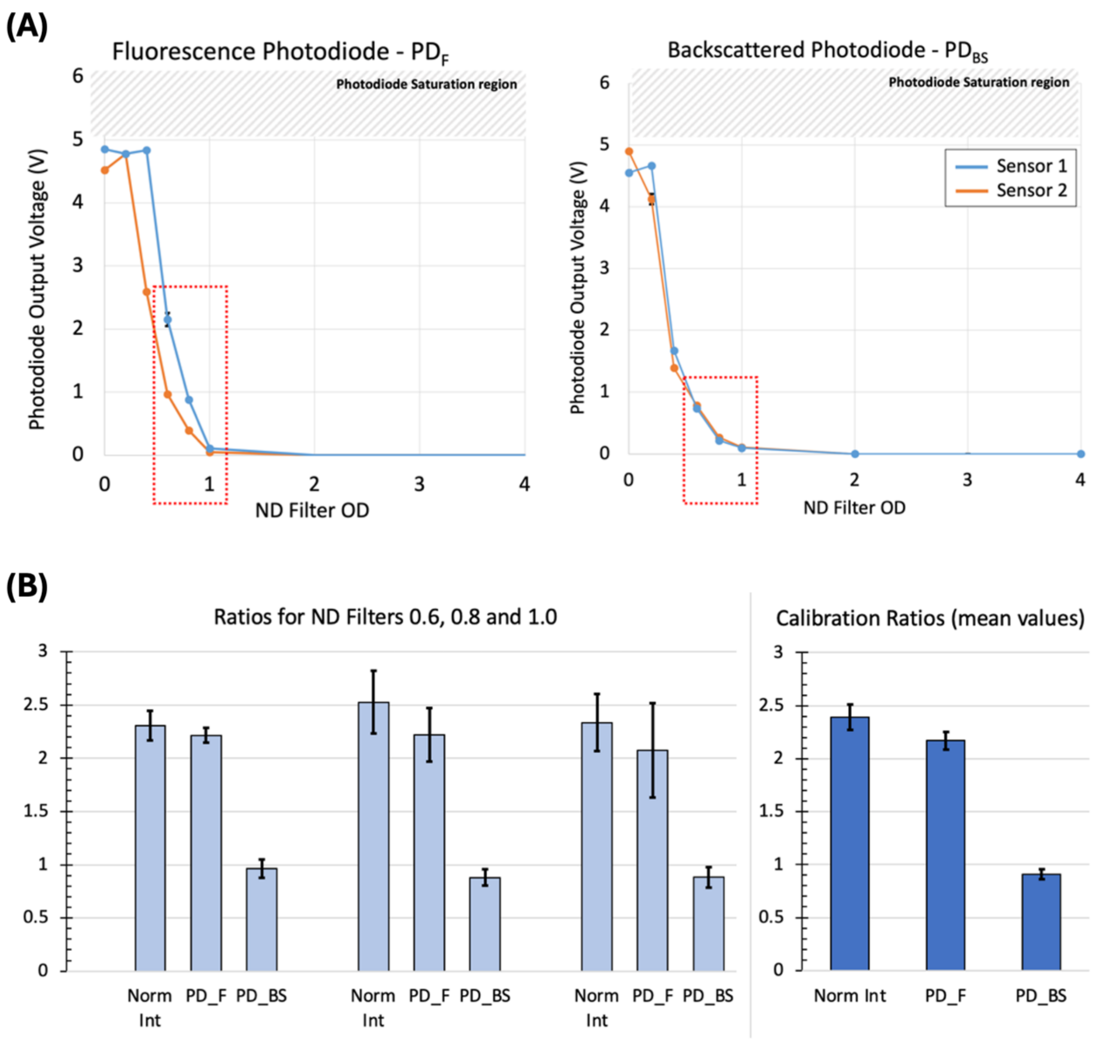


**Figure S2.** Calibration results. **(A)** Photodiode output signals from both sensors for the different ND filters used for calibration. Sensor 1 (UK) – blue; Sensor 2 (Zambia) – orange. Both photodiodes (fluorescence – left; backscatter – right) saturate at a voltage value of 5V due to the configuration of the electronic circuit. Values used to calculate the calibration parameters are highlighted with a dotted red box (chosen as they lie within the non-saturated and non-zero regions). These correspond to the values from the 0.6, 0.8 and 1.0 ND filters. **(B)** Mean sensor 1 : sensor 2 ratios (with standard deviation error bars) calculated for the fluorescence photodiode voltage (PD_F), backscatter photodiode voltage (PD_BS), and the normalized fluorescence intensity (Norm Int, calculated as described in [1]) for measurements using the 0.6, 0.8 and 1.0 ND filters. Values were calculated from measurements made on three different days with 10 repeat measurements made on each day. (Left) Individual ratios from each ND filter: 0.6 (left), 0.8 (middle), 1.0 (right). (right) Mean ratios averaged across the 0.6, 0.8 and 1.0 ND filter measurements. Values shown on this graph represent the final calibration ratios used for signal correction – i.e. the normalised intensity recorded by Sensor 1 was found to be 2.4x higher than that recorded by Sensor 2. Hence, Sensor 2 normalized intensity values were corrected by multiplying by 2.4 (the calibration ratio). Exact photodiode voltages and calibration ratios are shown in Table S3.

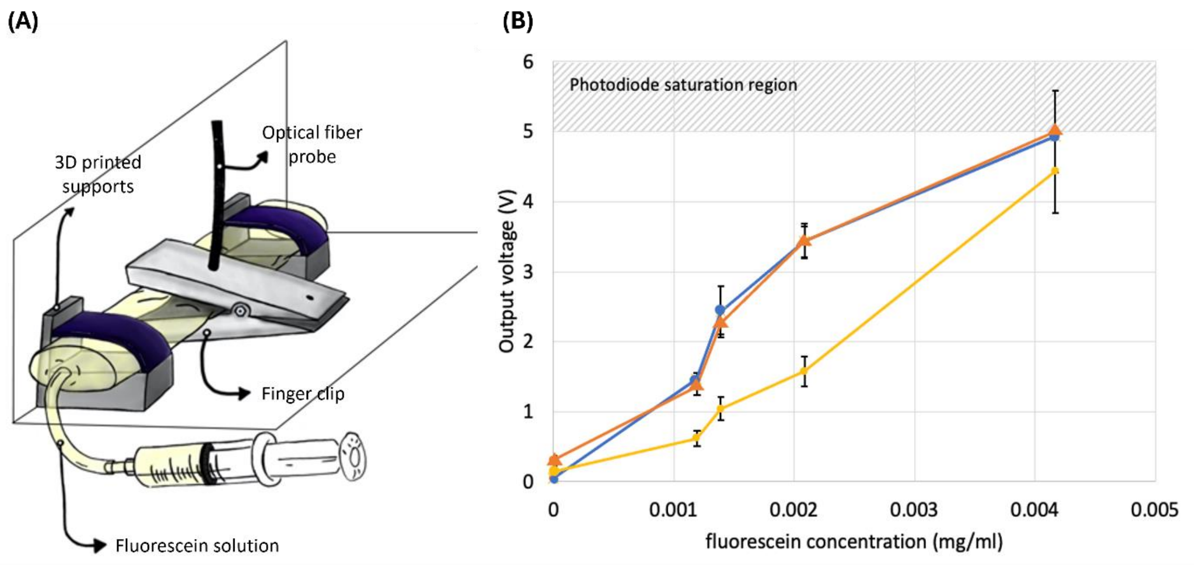


**Figure S3.** *In vitro* validation of sensor calibration. **(A)** Diagram of experimental setup. The optical fibre probe is clamped (using the 3D printed finger clip) in contact with a transparent thermoplastic container filled with aqueous fluorescein solutions of varying concentrations. The experimental setup is contained within a black box to block ambient light. The transparent container is attached to the inside of the box using 3D printed supports to maintain a constant position during measurements. **(B)** Output voltage detected by the fluorescence photodiodes for five different fluorescein concentrations. Blue curve indicates voltages measured with Sensor 1. Yellow curve indicates voltages measured with Sensor 2. Orange curve represents the corrected values for Sensor 2 (i.e. Sensor 2 output voltages multiplied by the fluorescence photodiode calibration ratio – see Table S3). The photodiode saturation region is shown (above 5V), demonstrating that the measurement made with Sensor 1 has reached saturation at the maximum fluorescein concentration. The corrected value for Sensor 2 is thus set to 5V at the maximum fluorescein concentration to avoid calculation of a saturated voltage value. Following calibration, Sensor 1 and Sensor 2 values are in good agreement (i.e. compare blue and orange curves).

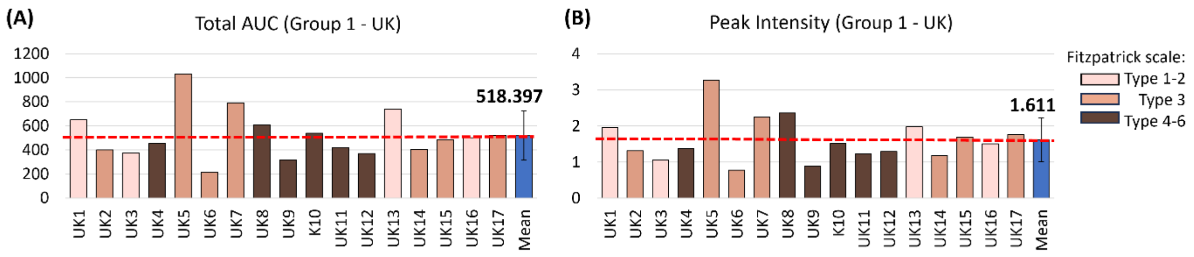


**Figure S4**. Total AUC and peak intensity values extracted from fluorescence signals recorded in all participants in Group 1 (UK). **(A)** Total AUC values measured in each participant. **(B)** Peak (maximum) intensity values measured in each participant. Bar colour indicates skin tone (assessed using the Fitzpatrick scale [2]; see Legend). Blue bars denote mean values (± standard deviation).

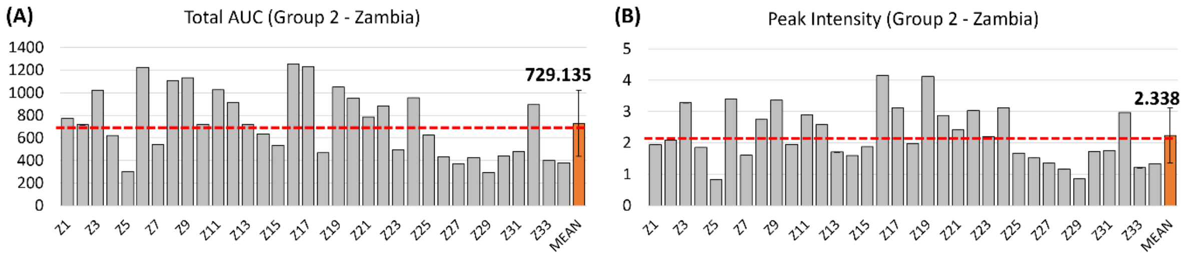


**Figure S5.** Total AUC and peak intensity values extracted from fluorescence signals recorded in all participants in Group 2 (Zambia). **(A)** Total AUC values measured in each participant. **(B)** Peak (maximum) intensity values measured in each participant. Grey bars represent individual participant values. Orange bars denote mean values (± standard deviation). Skin tone is not visualised in this figure as all participants in Group 2 were classified as Fitzpatrick type 5/6.

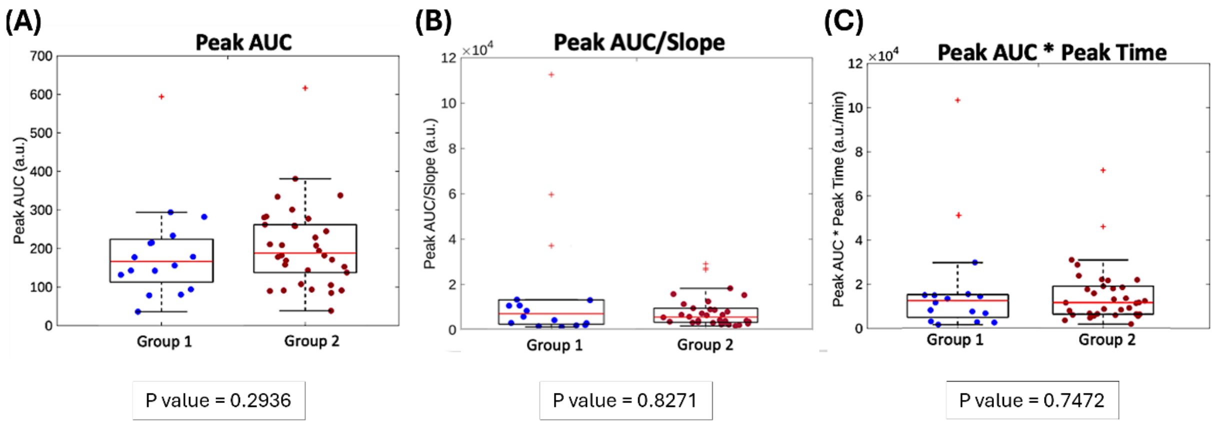


**Figure S6**. Scatter-box plots displaying the parameters extracted from the fluorescence vs. time curves for Group 1 (UK) and Group 2 (Zambia). **(A)** Peak AUC. **(B)** Peak AUC/Slope. **(C)** Peak AUC*Peak Time. Box plots show individual data points (circles), median values (red lines), 25th and 75th percentiles (upper and lower bounds of black box), most extreme data points excluding outliers (whiskers), and outliers (red crosses). Group 1 – UK (blue); Group 2 – Zambia (red). Outliers were defined as points that fell below the 25th percentile or above the 75th percentile by more than 1.5 times the interquartile range. Importantly, outliers are shown for visualization purposes only, and all data points (including outliers) were included in all calculations and analysis. Inset p-values represent results of Wilcoxon Rank Sum tests comparing Groups 1 and 2.

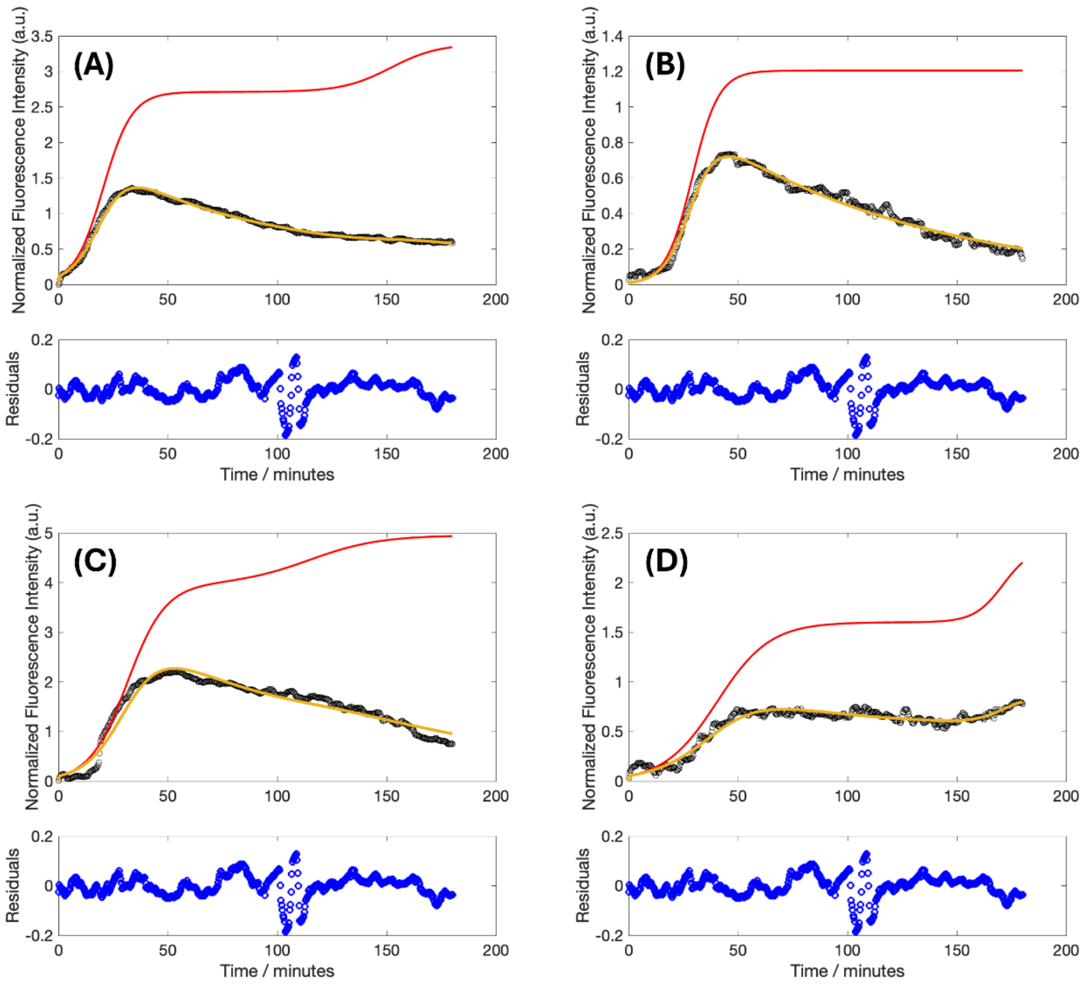


**Figure S7.** Example PBFK fits for Group 1 (UK). Black circles represent raw data, yellow lines represent fitted PBFK curves, red lines represent uptake-only curves (where effect of elimination has been removed). Subset graphs (blue circles) represent residuals (differences between fit and data at each time point). In all cases, the PBFK model is in close agreement with the data across the full time course and residuals appear randomly distributed around zero, indicating good fits. **(A)** Participant UK4. **(B)** Participant UK6. **(C)** Participant UK7. **(D)** Participant UK9.

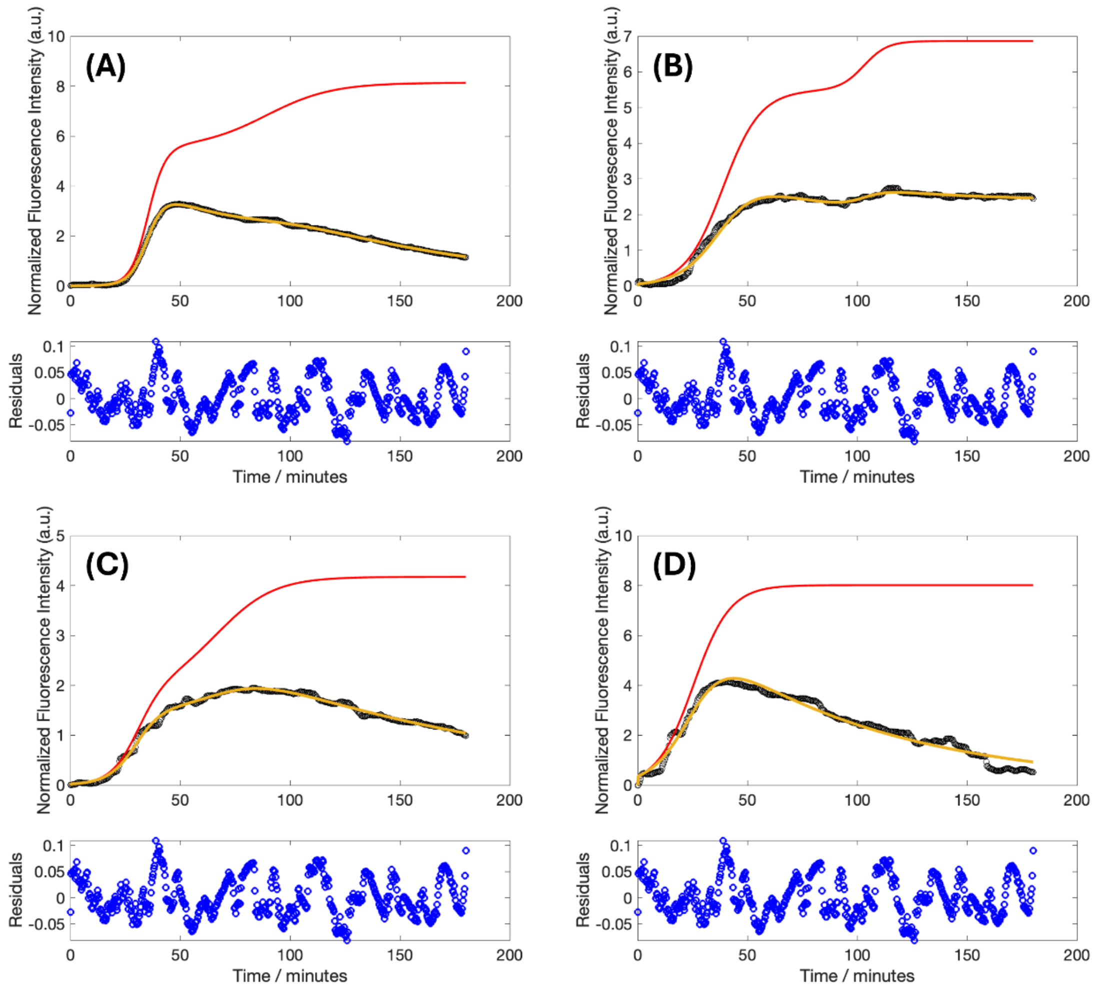


**Figure S8.** Example PBFK fits for Group 2 (Zambia). Black circles represent raw data, yellow lines represent fitted PBFK curves, red lines represent uptake-only curves (where effect of elimination has been removed). Subset graphs (blue circles) represent residuals (differences between fit and data at each time point). In all cases, the PBFK model is in close agreement with the data across the full time course and residuals appear randomly distributed around zero, indicating good fits. **(A)** Participant Z4. **(B)** Participant Z10. **(C)** Participant Z12. **(D)** Participant Z18.

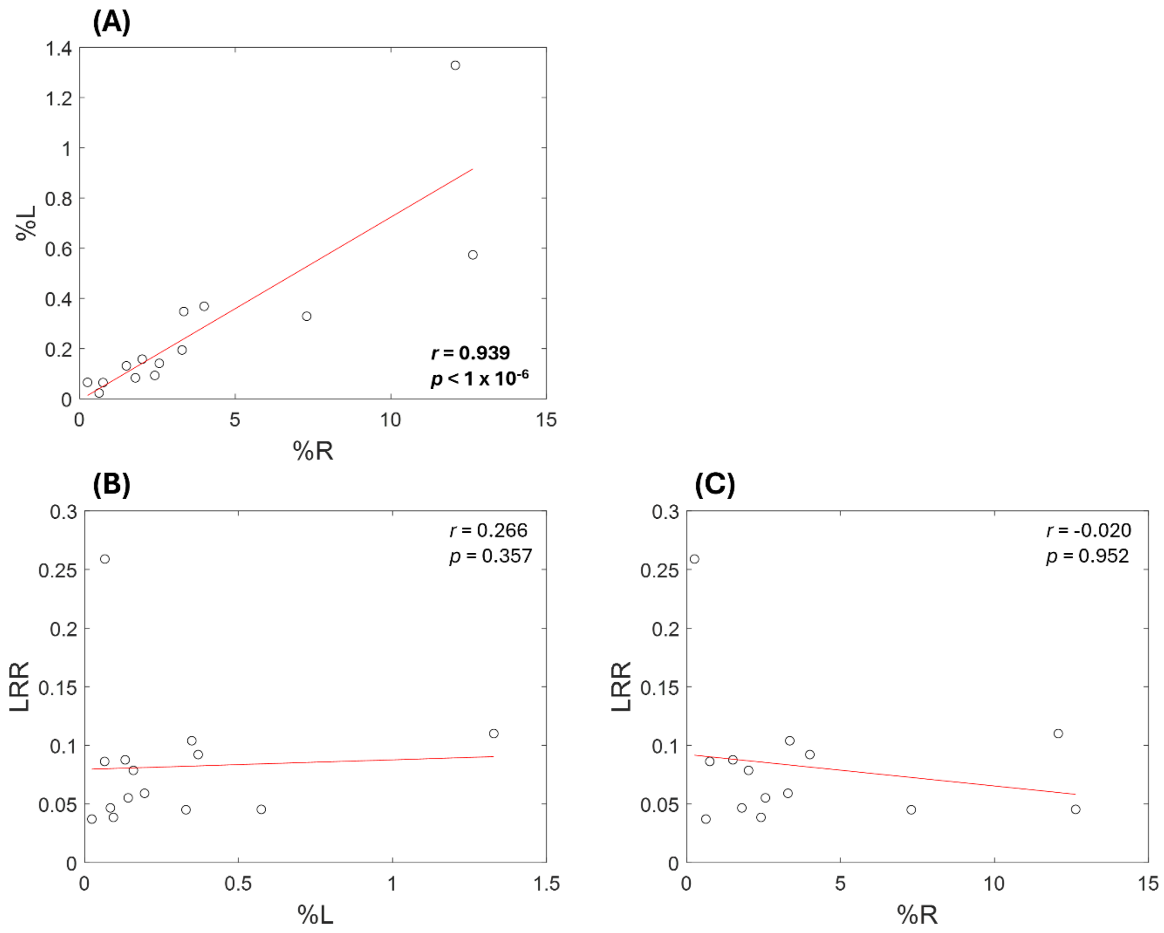


**Figure S9.** Correlations within LR parameters. **(A)** Percentage lactulose recovery (%L) vs. percentage rhamnose recovery (%R). **(B)** Lactulose:rhamnose ratio (LRR) vs. %L. **(C)** LRR vs. %R. Red lines represent linear regression trend lines. Inset values represent Spearman's Rank correlation coefficients (*r*) and corresponding *p* values. Significant correlation (between %L and %R) is shown in bold.

#### 2. Supplementary Tables

**Table S1.** Demographics of the study participants in Group 1 – UK. Lower region of table presents the percentage of female and male participants and the percentage (and number) of participants in each Fitzpatrick skin tone class [2]. BMI – body mass index. Yellow cells highlight participants with BMI>25. Red cell highlights participant with BMI>30.

| Group 1 - UK | | | | |
|---|---|---|---|---|
| Participant | Age | Sex | Fitzpatrick class | BMI |
| UK1 | 18-25 | F | 1 | - |
| UK2 | 26-30 | M | 2 | 24.8 |
| UK3 | 18-25 | F | 1 | - |
| UK4 | 36-40 | M | 5 | 30.8 |
| UK5 | 31-35 | F | 3 | 18.5 |
| UK6 | 18-25 | M | 3 | 23.5 |
| UK7 | 31-35 | M | 3 | 21.9 |
| UK8 | 26-30 | M | 5 | 24.1 |
| UK9 | 31-35 | M | 6 | 24.7 |
| UK10 | 26-30 | M | 5 | - |
| UK11 | 18-25 | M | 5 | 22.1 |
| UK12 | 18-25 | M | 5 | 19.8 |
| UK13 | 36-40 | F | 2 | 19.7 |
| UK14 | 61-65 | M | 3 | 27.4 |
| UK15 | 36-40 | M | 3 | 25.2 |
| UK16 | 31-35 | M | 2 | - |
| UK17 | 36-40 | M | 3 | 28.1 |
| Female / Male | | | 23.5% / 76.5% | |
| Fitzpatrick class (1-2) | | | 29.4% (5) | |
| Fitzpatrick class (3-4) | | | 35.3% (6) | |
| Fitzpatrick class (5-6) | | | 35.3% (6) | |

**Table S2.** Demographics of the study participants in Group 2 – Zambia. Lower region of table presents the percentage of female and male participants and the percentage (and number) of participants in each Fitzpatrick skin tone class [2]. BMI – body mass index. Yellow cells highlight participants with BMI>25. Red cells highlight participants with BMI>30.

| Group 2 - Zambia | | | | | | | |
|---|---|---|---|---|---|---|---|
| Participant | Age | Sex | BMI | Participant | Age | Sex | BMI |
| **Z1** | 31-35 | F | 24.20 | **Z18** | 31-35 | F | 29.2 |
| **Z2** | 18-25 | M | 20.5 | **Z19** | 18-25 | M | 20.7 |
| **Z3** | 41-45 | F | 25.8 | **Z20** | 41-45 | F | 26.7 |
| **Z4** | 26-30 | F | 20 | **Z21** | 31-35 | F | 31.3 |
| **Z5** | 31-35 | M | 25.6 | **Z22** | 18-25 | M | 19.1 |
| **Z6** | 31-35 | F | 20.7 | **Z23** | 18-25 | F | 25.3 |
| **Z7** | 51-55 | M | 21.5 | **Z24** | 26-30 | F | 26.9 |
| **Z8** | 56-60 | M | 26.3 | **Z25** | 18-25 | M | 21.1 |
| **Z9** | 26-30 | M | 25.2 | **Z26** | 26-30 | F | 32.4 |
| **Z10** | 26-30 | F | 28.4 | **Z27** | 36-40 | F | 21.8 |
| **Z11** | 18-25 | M | 24.6 | **Z28** | 18-25 | M | 20.8 |
| **Z12** | 18-25 | F | 28.8 | **Z29** | 18-25 | M | 19.9 |
| **Z13** | 26-30 | F | 26.3 | **Z30** | 18-25 | M | 23.1 |
| **Z14** | 31-35 | F | 20.6 | **Z31** | 46-50 | F | 25.2 |
| **Z15** | 46-50 | M | 21.9 | **Z32** | 36-40 | F | 39.7 |
| **Z16** | 51-55 | M | 19.8 | **Z33** | 26-30 | F | 38.6 |
| **Z17** | 26-30 | F | 23.2 | **Z34** | 18-25 | M | 20.2 |
| Female / Male | | | | 55.9% / 44.1% | | | |
| Fitzpatrick class (1-2) | | | | 0 | | | |
| Fitzpatrick class (3-4) | | | | 0 | | | |
| Fitzpatrick class (5-6) | | | | 100% (34) | | | |

**Table S3.** Calibration output voltages (recorded using Sensor 1 and 2) and resulting calibration ratios. Only values recorded using the 0.6, 0.8 and 1.0 ND filters were used for calculation of calibration ratios for the normalized intensity. Lower and higher values – corresponding to the saturation or zero regions – are not shown (as these introduce errors into the calculation of the calibration ratios). Mean calibration ratios were calculated by averaging the calibration ratios from the 0.6, 0.8 and 1.0 ND filters. These values were then used to correct the data collected with Sensor 2 to allow fair comparison of data collected in Groups 1 and 2. $PD_F$ – fluorescence photodiode; $PD_{BS}$ – backscatter photodiode; OD – optical density; STD – standard deviation.

| Output Voltage | | Filter OD | | | | | | | | |
|---|---|---|---|---|---|---|---|---|---|---|
| | | 0 | 0.2 | 0.4 | 0.6 | 0.8 | 1.0 | 2.0 | 3.0 | 4.0 |
| **$PD_F$** | Sensor 1 | 4.715 | 4.703 | 4.646 | 2.184 | 1.404 | 0.096 | $-1.002*10^{-3}$ | $-1.352*10^{-4}$ | $-1.497*10^{-4}$ |
| | Sensor 2 | 4.634 | 4.711 | 2.569 | 0.984 | 0.625 | 0.046 | $5.751*10^{-3}$ | $5.640*10^{-3}$ | $2.491*10^{-3}$ |
| **$PD_{BS}$** | Sensor 1 | 4.549 | 4.599 | 1.752 | 0.753 | 0.529 | 0.091 | $-2.202*10^{-5}$ | $-1.189*10^{-5}$ | $-2.956*10^{-6}$ |
| | Sensor 2 | 4.656 | 4.372 | 1.374 | 0.781 | 0.615 | 0.104 | $-5.735*10^{-5}$ | $-5.100*10^{-5}$ | $1.125*10^{-3}$ |

| Calibration Ratio | Filter OD | | | | | | | | |
|---|---|---|---|---|---|---|---|---|---|
| | 0 | 0.2 | 0.4 | 0.6 | 0.8 | 1.0 | 2.0 | 3.0 | 4.0 |
| **$PD_F$** | 1.018 | 0.998 | 1.807 | 2.216 | 2.244 | 2.075 | 530.926 | 5.591 | 9.119 |
| **$PD_{BS}$** | 0.978 | 1.054 | 1.276 | 0.963 | 0.880 | 0.881 | 0.932 | 1.464 | 0.071 |
| **Normalised ($PD_F$ / $PD_{BS}$)** | | | | 2.309 | 2.558 | 2.335 | | | |

| Mean calibration Ratio | Mean | STD |
|---|---|---|
| **$PD_F$** | 2.178 | 0.091 |
| **$PD_{BS}$** | 0.908 | 0.047 |
| **Normalised ($PD_F$ / $PD_{BS}$)** | 2.401 | 0.137 |

**Table S4.** TFS parameters measured in each participant in Group 1. Mean and standard deviation (STD) values are shown in the lowest two rows of the table. Means and standard deviations were calculated by averaging the values for each individual (i.e. not by extracting parameters from the mean fluorescence vs. time curve shown in Fig. 3B). Values for Participant UK8 are shown for completeness only. Values for participant UK8 were excluded from all analysis (including calculation of mean values) due to an aberrant fluorescence signal resulting from errors during data collection.

| Participant | Total AUC (a.u.) | Peak Intensity (a.u.) | Peak Time (min) | Peak AUC (a.u.) | Peak AUC/Slope (a.u.*min) | Peak AUC * Peak Time (a.u.*min) |
|---|---|---|---|---|---|---|
| UK1 | 647.962 | 1.956 | 173 | 594.885 | 58595.454 | 103113.570 |
| UK2 | 399.798 | 1.311 | 74 | 179.018 | 10340.847 | 13247.401 |
| UK3 | 371.823 | 1.057 | 126 | 233.879 | 36373.363 | 29624.749 |
| UK4 | 452.754 | 1.377 | 34 | 78.758 | 1765.022 | 2677.798 |
| UK5 | 1031.323 | 3.271 | 52 | 282.719 | 4042.835 | 14795.679 |
| UK6 | 215.303 | 0.775 | 41 | 36.568 | 1886.961 | 1511.502 |
| UK7 | 788.615 | 2.251 | 47 | 156.531 | 2739.857 | 7461.292 |
| UK8 | 607.884 | 2.356 | 44 | 200.932 | 7232.329 | 8974.992 |
| UK9 | 317.420 | 0.892 | 174 | 294.702 | 110715.817 | 51278.258 |
| UK10 | 537.911 | 1.511 | 71 | 215.720 | 10407.111 | 15316.135 |
| UK11 | 420.390 | 1.225 | 80 | 143.426 | 8086.627 | 11521.901 |
| UK12 | 370.996 | 1.288 | 61 | 132.324 | 5571.689 | 8071.811 |
| UK13 | 741.229 | 1.973 | 46 | 142.771 | 2821.515 | 6662.643 |
| UK14 | 404.873 | 1.181 | 80 | 177.666 | 12980.043 | 14272.545 |
| UK15 | 484.334 | 1.687 | 30 | 80.912 | 1149.392 | 2481.313 |
| UK16 | 501.114 | 1.504 | 69 | 214.303 | 12740.416 | 14858.376 |
| UK17 | 519.023 | 1.757 | 31 | 94.578 | 1407.103 | 2963.466 |
| **Mean** | **512.804** | **1.563** | **74.625** | **191.173** | **17601.503** | **18741.152** |
| **STD** | **203.343** | **0.607** | **45.599** | **129.791** | **29145.270** | **25684.797** |

**Table S5**. TFS and LR parameters measured in each participant in Group 2. Mean and standard deviation (STD) values are shown in the lowest two rows of the table. Means and standard deviations of TFS parameters were calculated by averaging the values for each individual (i.e. not by extracting parameters from the mean fluorescence vs. time curve shown in Fig. 3B). For LR analysis, null values (labelled as "-") correspond to participants who did not take an LR test.

| ID | Total AUC (a.u.) | Peak Intensity (a.u.) | Peak Time (min) | Peak AUC (a.u.) | Peak AUC/Slope (a.u.*min) | Peak AUC* Peak Time (a.u.*min) | L (%) | R (%) | LRR |
|---|---|---|---|---|---|---|---|---|---|
| Z1 | 771.566 | 2.086 | 95 | 390.733 | 15002.000 | 28737.674 | 0.065 | 0.253 | 0.259 |
| Z2 | 715.383 | 2.149 | 40 | 148.512 | 2988.800 | 6299.497 | 0.132 | 1.500 | 0.088 |
| Z3 | 1020.342 | 3.291 | 48 | 145.041 | 2380.800 | 7837.068 | 1.329 | 12.070 | 0.110 |
| Z4 | 622.175 | 1.895 | 38 | 93.194 | 1955.000 | 3493.954 | 0.065 | 0.750 | 0.086 |
| Z5 | 302.671 | 1.052 | 119 | 185.698 | 26185.000 | 21575.149 | 0.023 | 0.623 | 0.037 |
| Z6 | 1222.488 | 3.491 | 50 | 248.846 | 3943.700 | 13463.278 | - | - | - |
| Z7 | 540.058 | 1.678 | 77 | 182.678 | 10997.000 | 17555.826 | - | - | - |
| Z8 | 1105.918 | 2.901 | 118 | 635.637 | 26855.000 | 71664.989 | - | - | - |
| Z9 | 1134.237 | 3.463 | 37 | 181.413 | 2004.000 | 6643.987 | - | - | - |
| Z10 | 722.143 | 2.111 | 91 | 327.087 | 12182.000 | 23745.884 | - | - | - |
| Z11 | 1026.211 | 3.019 | 73 | 409.241 | 2996.100 | 8606.254 | - | - | - |
| Z12 | 913.892 | 2.675 | 72 | 298.479 | 8459.800 | 21863.586 | - | - | - |
| Z13 | 717.742 | 1.738 | 54 | 131.737 | 7676.600 | 12996.513 | - | - | - |
| Z14 | 636.684 | 1.646 | 131 | 432.528 | 28771.000 | 46064.785 | - | - | - |
| Z15 | 534.301 | 1.986 | 86 | 268.846 | 8653.000 | 15752.383 | - | - | - |
| Z16 | 1256.022 | 4.353 | 40 | 280.905 | 2745.500 | 11492.530 | - | - | - |
| Z17 | 1231.354 | 3.296 | 47 | 258.695 | 6063.300 | 19008.895 | - | - | - |
| Z18 | 468.528 | 2.042 | 61 | 109.023 | 3252.700 | 6429.938 | - | - | - |
| Z19 | 1049.534 | 4.193 | 46 | 199.939 | 2968.800 | 12311.042 | - | - | - |
| Z20 | 951.076 | 3.041 | 53 | 175.521 | 3254.700 | 9107.232 | - | - | - |
| Z21 | 784.373 | 2.501 | 79 | 220.378 | 9194.500 | 22028.587 | - | - | - |
| Z22 | 882.168 | 3.171 | 49 | 234.035 | 3692.200 | 11261.964 | - | - | - |
| Z23 | 495.642 | 2.239 | 62 | 84.598 | 2865.400 | 5950.252 | - | - | - |
| Z24 | 955.296 | 3.174 | 38 | 143.437 | 1744.200 | 5494.868 | - | - | - |
| Z25 | 624.717 | 1.752 | 83 | 243.585 | 6892.100 | 11582.493 | - | - | - |
| Z26 | 434.615 | 1.575 | 60 | 94.171 | 3711.900 | 5431.818 | 0.093 | 2.414 | 0.039 |
| Z27 | 368.945 | 1.391 | 86 | 145.817 | 6295.500 | 8272.372 | 0.194 | 3.289 | 0.059 |
| Z28 | 423.062 | 1.212 | 86 | 212.116 | 15434.000 | 17948.346 | 0.141 | 2.558 | 0.055 |
| Z29 | 291.321 | 0.931 | 79 | 140.771 | 5431.400 | 4620.971 | 0.084 | 1.791 | 0.047 |
| Z30 | 438.035 | 1.921 | 36 | 94.759 | 3604.000 | 6237.123 | 0.369 | 4.000 | 0.092 |
| Z31 | 477.527 | 1.786 | 118 | 265.363 | 17974.000 | 30956.536 | 0.348 | 3.347 | 0.104 |
| Z32 | 895.588 | 3.125 | 65 | 265.888 | 6276.200 | 18472.876 | 0.329 | 7.295 | 0.045 |
| Z33 | 399.902 | 1.218 | 66 | 94.574 | 5149.200 | 5832.684 | 0.158 | 2.010 | 0.079 |
| Z34 | 377.082 | 1.393 | 46 | 41.253 | 1381.100 | 1774.232 | 0.574 | 12.635 | 0.045 |
| **MEAN** | **729.135** | **2.338** | **68** | **217.191** | **7911.19118** | **15309.282** | **0.279** | **3.895** | **0.082** |
| **STD** | **293.207** | **0.896** | **26.376** | **120.877** | **7405.14798** | **13660.981** | **0.339** | **3.991** | **0.057** |

**Table S6.** Demographic analysis between groups – Group 1 (UK) vs. Group 2 (Zambia). Table shows mean peak intensity values extracted from the subgroups presented in Fig. 4. Bottom row shows *p* values from Wilcoxon Rank Sum tests comparing Group 1 and Group 2 within each demographic category. Significant differences ($p < 0.05$) are highlighted in green.

| Parameters | SEX | | AGE | | BMI | |
|---|---|---|---|---|---|---|
| | Female | Male | < 35 years | > 35 years | < 25 | > 25 |
| **Mean Group 1** | 2.064 | 1.397 | 1.615 | 1.478 | 1.623 | 1.501 |
| **Mean Group 2** | 2.332 | 2.335 | 2.227 | 2.605 | 2.309 | 2.371 |
| ***p value*** | 0.543 | 0.021 | 0.067 | 0.011 | 0.040 | 0.042 |

**Table S7.** Wilcoxon Rank Sum test results from within-group comparisons across demographics.

| *P-values* from Wilcoxon Rank Sum tests | | | |
|---|---|---|---|
| Parameters | SEX | AGE | BMI |
| | Female vs Male | < 35 vs > 35 years | < 25 vs > 25 value |
| **Group 1 *p value*** | 0.1703 | 1 | 0.9527 |
| **Group 2 *p value*** | 0.7287 | 0.3744 | 0.6414 |

**Table S8.** Correlation of LR and TFS parameters. Spearman's Rank correlation coefficients (*r,* top) and *p*-values (bottom) for correlations between TFS parameters (Total AUC, Peak Intensity, Peak Time, Peak AUC, Peak AUC/Slope and Peak AUC*Peak Time) and LR parameters (%L, %R and LRR). Significant correlations ($p < 0.05$) are highlighted in green.

| Spearman's Rank correlation coefficient (*r*) | | | | | | |
|---|---|---|---|---|---|---|
| Parameters | Total AUC | Peak Int | Peak Time | Peak AUC | Peak AUC/Slope | Peak AUC* Peak Time |
| **%L** | 0.2527 | 0.3802 | -0.3102 | -0.1077 | -0.3187 | -0.0286 |
| **%R** | 0.1473 | 0.2703 | -0.3080 | -0.1780 | -0.3099 | -0.1165 |
| **LRR** | 0.5780 | 0.5473 | -0.1364 | 0.2176 | -0.1341 | 0.2440 |

| *P-values* from Spearman's Rank correlation analysis | | | | | | |
|---|---|---|---|---|---|---|
| Parameters | Total AUC | Peak Int | Peak Time | Peak AUC | Peak AUC/Slope | Peak AUC* Peak Time |
| **%L** | 0.3825 | 0.1808 | 0.2804 | 0.7156 | 0.2665 | 0.9276 |
| **%R** | 0.6158 | 0.3491 | 0.2840 | 0.5423 | 0.2806 | 0.6930 |
| **LRR** | 0.0334 | 0.0459 | 0.6419 | 0.4541 | 0.6485 | 0.3998 |

### 3. Supplementary Methods

#### 3.1. Lactulose:rhamnose (LR) testing

For LR testing, participants provided a baseline urine sample and then completely voided their bladders before ingesting the test solution. All urine was then collected up to 3 hours post-ingestion and the total volume of urine collected was measured. Baseline and LR urine samples were aliquoted and stored in a -80°C freezer. Aliquots were later shipped from Zambia to the UK (on dry ice) for analysis, where concentrations of lactulose and rhamnose were measured using liquid chromatography – tandem mass spectrometry (LC-MS/MS). Full details of the LC-MS/MS method and the calculation of percentage recoveries of lactulose and rhamnose (and the LR ratio, LRR) are provided below.

#### 3.2. Measurement of lactulose and rhamnose concentrations in urine samples using liquid chromatography – tandem mass spectrometry

Lactulose and rhamnose concentrations in urine samples were measured using our previously reported targeted liquid chromatography – tandem mass spectrometry (LC-MS/MS) method [3]. The method was fully validated according to the established guidelines for bioanalytical method validation (see "M10 Bioanalytical Method Validation and Study Sample Analysis," available at https://www.fda.gov/media/162903/download).

All instrumental and experimental parameters, LC-MS/MS setup, solvents and chromatographic column were as reported for lactulose:mannitol assay [3]. The information specific for the lactulose:rhamnose assay and a brief description of the LC-MS/MS analysis are detailed below.

##### 3.2.1. Chemicals and materials

Lactulose and rhamnose analytical standards were purchased from Sigma-Aldrich (Gillingham, UK) and Fluorochem Ltd. (Derbyshire, UK). Stable isotope labelled internal standard of lactulose-$^{13}C_{12}$ was purchased from Merck Life Science (Gillingham, UK), and L-[UL-$^{13}C_6$]-rhamnose monohydrate was purchased from Omicron Biochemicals, Inc. (South Bend IN, USA).

##### 3.2.2. LC-MS/MS method and data processing

The method was developed and set up for the analysis of urinary sugars on an ACQUITY UPLC solvent management system using a 2777C external autosampler (Waters, Wilmslow, UK) coupled with a Xevo TQ-S tandem quadrupole instrument (Waters, Wilmslow, UK) with electrospray ionization (ESI) in negative ion mode. The LC column used to separate sugars was an ACQUITY UPLC BEH Amide 1.7 µm, 2.1 × 150 mm column (Waters Corporation) thermostatted at 60°C. The LC gradient [3] was performed at 0.4 ml/min using 5 mM ammonium acetate (pH 9.3) in water as mobile phase A and 10% methanol in acetonitrile (v/v) as mobile phase B. The injection volume was 2 µL.

Lactulose and rhamnose along with their stable isotope labelled standards were measured using multiple reaction monitoring (MRM) transitions presented in **Table S9**. The desolvation gas was nitrogen, and the collision gas was argon. All MS detection parameters were specified in [3].

Raw LC-MS/MS spectral data were visualized using MassLynx (v4.1) software (Waters Corporation). Peak integration, compound quantification and quality control were carried out using the TargetLynx application package within the MassLynx (v4.1) software.

**Table S9.** MS/MS transitions used for measuring lactulose, rhamnose and their stable isotope labelled internal standards.

| Analyte | Parent *m/z* | Quantifier *m/z* | Qualifier *m/z* | Collision Energy, eV | Cone Voltage, V |
|---|---|---|---|---|---|
| Lactulose | **341.17** | **160.88** | | 6 | 22 |
| | 341.17 | | 100.82 | 14 | 22 |
| | 341.17 | | 178.82 | 8 | 22 |
| Lactulose-$^{13}C_{12}$ | **353.15** | **166.86** | | 6 | 20 |
| | 353.15 | | 104.86 | 14 | 20 |
| | 353.15 | | 184.87 | 6 | 20 |
| Rhamnose | **163.09** | **58.84** | | 12 | 44 |
| | 163.09 | | 88.86 | 6 | 44 |
| | 163.09 | | 102.84 | 6 | 44 |
| Rhamnose-$^{13}C_6$ | **169.08** | **60.85** | | 14 | 44 |
| | 169.08 | | 91.86 | 10 | 44 |
| | 169.08 | | 106.87 | 8 | 44 |

##### 3.2.3. LC-MS/MS analysis of urinary lactulose and rhamnose

Individually prepared stock solutions of lactulose and rhamnose were mixed at different concentration levels to generate a series of calibration solutions within the range of 20 µg/ml (lower limit of quantification, LLOQ) to 1000 µg/ml (upper limit of quantification, ULOQ) for both sugar probes. The internal standards mixture contained 50 µg/ml of lactulose-$^{13}C_{12}$ and 100 µg/ml of rhamnose-$^{13}C$. Quality control (QC) samples used to monitor accuracy and precision of the quantitative assay were prepared at concentrations of 20 µg/ml (LLOQ QC), 100 µg/ml, 250 µg/ml, 500 µg/ml, and 750 µg/ml (high QC).

Study urine samples, calibration and QC solutions were prepared as detailed in [3]. Urine samples were preconcentrated four times by drying 80 µl of each sample under a gentle stream of nitrogen at room temperature followed by reconstitution in 20 µl of LC-MS grade water. Then, 20 µl of the stable isotope labelled internal standards mix and 120 µl of cold 1:2 methanol:acetonitrile mixture were added to each reconstituted sample. Samples were vortex mixed after each addition, and then the plates with the samples were sealed and mixed at 1400 rpm for two minutes at 4°C (MixMate, Eppendorf). Finally, the plates were centrifuged for ten minutes at 3486g and 4°C prior to the LC-MS/MS analysis.

Blocks of 40 urine samples, collected from each patient at baseline and after sugar intake, were analyzed in randomized order and quantified using a nine-point calibration curve run along with the study samples and three full sets of QC samples to monitor accuracy and precision of the analysis.

#### 3.3. Calculation of percentage urinary recoveries of lactulose and rhamnose and LRR

The percentage recovery of lactulose and rhamnose in urine (%L and %R respectively) were calculated using equations (S1) and (S2).

$$\%L = 100 \times \left(V \times \frac{C_L}{D_L}\right) \tag{S1}$$

$$\%R = 100 \times \left(V \times \frac{C_R}{D_R}\right) \tag{S2}$$

In these equations, $V$ represents the total volume of urine collected, $C_L$ and $C_R$ represent the urine concentrations of lactulose and rhamnose respectively, and $D_L$ and $D_R$ represent the administered doses of lactulose and rhamnose respectively (5 g for lactulose and 1 g for rhamnose). The LR ratio (LRR) was then calculated according to equation (S3).

$$LRR = \frac{\%L}{\%R} \tag{S3}$$

#### 3.4. Data and statistical analysis

##### 3.4.1. Fluorescence signal analysis

Normalized fluorescence signals (calculated as described in [1]) were first manually observed for each group as both individual time course plots and as mean ± standard deviation time course graphs (**Fig. 3A&B**; **Fig. S4&S5,** Supplementary Information). To facilitate statistical comparison of the two groups, a series of parameters were then extracted from the normalized fluorescence signals including: peak (maximum) intensity; total area under the curve (AUC); peak time; peak AUC (i.e. AUC from the start of the measurement up to the peak time); peak AUC divided by the average gradient of the graph in the uptake region (i.e. from the start of the measurement up to the peak time; peak AUC / slope); and peak AUC multiplied by peak time (peak AUC * peak time). These parameters were chosen as previous work demonstrated that they provide detection of changes in intestinal permeability [4] and gastric emptying rate [5]. Datasets were smoothed using a median filter (window width = 10) and then baseline subtracted prior to calculation of these parameters to reduce the impact of noise. Wilcoxon Rank Sum tests were then used to analyze differences between Groups 1 (UK) and 2 (Zambia).

##### 3.4.2. Fluorescence vs. LR correlation analysis

In the subset of Group 2 participants who took both a TFS and LR test, the percentage urinary recoveries of Lactulose and Rhamnose (i.e. %L and %R) and the Lactulose:Rhamnose ratio (LRR) were compared against the extracted fluorescence parameters. Scatter plots allowed visual observation of trends and Spearman's Rank correlation coefficients (*r*) were used to measure the correlations between fluorescence parameters and LR values (i.e. %L, %R and LRR).

##### 3.4.3. Physiologically based fluorescence kinetics modelling

As the collected fluorescence time courses exhibited considerable variability between participants and groups (see **Fig. 3A&B**), there was a limitation to the value of calculating individual parameters such as those proposed in section 2.4.1 (as these parameters inherently assess just a single component of the complex curves). Thus, we also analysed TFS data using a physiologically based fluorescence kinetics (PBFK) model developed in-house. This model was designed empirically based on visual observations of fluorescence vs. time curves and represents a similar approach to pharmacokinetic analysis used in drug uptake studies (e.g. see [6]).

Our in-house PBFK model built on the approach reported in [5] and incorporated two uptake pathways to model the permeation of fluorescein from the gut into the blood stream and two exponential decay pathways to model the elimination of fluorescein from the blood stream (via the kidneys and liver). This 4-compartment approach was chosen due to the shape of the observed fluorescence vs. time curves (see **Fig. 3A&B**): curves rise to a peak point before gradually decreasing back towards zero; some curves exhibit single peaks while others exhibit two peaks (necessitating two uptake pathways in the kinetic model); decay regions of curves are typically complex, exhibiting two or more decay rates (indicating the need for a minimum of two decay/elimination pathways in the kinetic model). Importantly, we note that the use of two uptake pathways in the kinetic model does not indicate two different routes of fluorescein absorption. Rather, it accounts for cyclical gastric emptying whereby fluorescein empties from the stomach into the intestine in multiple stages. This leads to observation of multiple peaks in some fluorescence datasets and necessitates use of two uptake pathways in the kinetic model. The PBFK model is mathematically described by the following formula.

$$I(t) = \underbrace{\left(ae^{-bt} + (1-a)e^{-ct}\right)}_{\text{Decay (elimination) pathways}} \underbrace{\left(\frac{B_{max}}{1 + e^{-k_B(t - t_{B1/2})}} + \frac{L_{max}}{1 + e^{-k_L(t - t_{L1/2})}}\right)}_{\text{Uptake (permeation) pathways}} \qquad \text{(S4)}$$

In equation (S4): *I*(*t*) represents the fluorescence intensity at time, *t*; *a* represents the relative contributions of the two decay (elimination) pathways; *b* and *c* represent the respective decay rates of the two elimination pathways (i.e. the rates at which fluorescein is eliminated from the blood stream); $B_{max}$ and $L_{max}$ represent the maximum intensity contributions (to the fluorescence signal) of the first and second uptake pathways respectively; $k_B$ and $k_L$ represent the logistic growth rates (steepness) of the two uptake pathways; and $t_{B½}$ and $t_{L½}$ represent the half-maximum times of the two uptake pathways (i.e. the time at which half of the maximum point would be reached in a single sigmoid uptake model).

For each dataset, equation (S4) was fitted to the observed fluorescence vs. time curve to extract nine model parameters (i.e. *a*, *b*, *c*, $B_{max}$, $L_{max}$, $k_B$, $k_L$, $t_{B½}$ and $t_{L½}$) which provided a complete description of the observed fluorescence kinetics. Fitting was achieved using the "*lsqcurvefit*" function in MATLAB. Models with single uptake and decay pathways were also tested and these provided reliably worse fits than the 4-compartment model presented in equation (S4).

Fitted parameters were then used to further explore trends in the data. First, "uptake-only" curves were plotted for each group by generating datasets based on the uptake parameters of the fit only (i.e. the parameters indicated as "Uptake (permeation) parameters" in equation (S4)). This removed the effect of elimination and provided an estimate of the total amount of fluorescein that had permeated from the gut into the blood stream.

The uptake-only curves and the final fluorescence intensities in those curves (which provided an indication of the total amount of fluorescein that had permeated the gut barrier) were first compared across Groups 1 and 2. Second, the fitted parameters were used in a regression analysis to investigate prediction of LR values using TFS data (see section 2.4.4). We note that the final fluorescence intensity in the uptake-only curves was measured at 150 minutes. This time point was chosen for two reasons: (1) plateaus had typically been observed by this stage of the experiments; (2) measurement at this time point avoided any errors introduced at the end of the datasets due to early removal of the fluorescence sensor.

##### 3.4.4. Regression analysis for TFS-based prediction of LR values

To explore TFS-based prediction of LR parameters, the fitted parameters extracted from fitting of equation (S4) were used in linear regression models to determine the relationship between the LR parameters and the fitted fluorescence values. The fluorescence parameters used in regression modelling included all fitted values from the "Uptake (permeation) pathways" component of equation (1) (i.e. $B_{max}$, $L_{max}$, $k_B$, $k_L$, $t_{B½}$ and $t_{L½}$) plus the final (maximum) fluorescence intensity observed in the "uptake-only" curves (hereafter referred to as $F_{final}$). These seven parameters were used to generate linear regression models according to

$$\%L, \%R, LRR = \beta_0 + \sum_{i=1}^{n} \beta_i x_i + \varepsilon \qquad \text{(S5)}$$

where %L, %R and LRR represent the dependent variables, $x_i$ are the independent variables representing the fitted fluorescence parameters (i.e. $B_{max}$, $L_{max}$, $k_B$, $k_L$, $t_{B½}$, $t_{L½}$ and $F_{final}$), $\beta_i$ are the regression coefficients, $\beta_0$ is the intercept, and $\varepsilon$ is an error term.

Three linear regression models were generated to train the fluorescence parameters on the %L, %R and LRR values respectively. Regression modelling was performed using MATLAB, with regression model fits achieved using the "*fitlm*" function. In all cases, model performance was evaluated using $R^2$ goodness-of-fit parameters, Spearman's Rank correlation coefficients ($r$; to assess the correlation between predicted and actual %L / %R / LRR values), and mean absolute error (MAE) values (between predicted and actual %L / %R / LRR values).

### 4. Supplementary Notes

#### 4.1. Supplementary Note 1: Peak time observations

When exploring the time at which peak intensity was reached in Groups 1 (UK) and 2 (Zambia), we observed that the mean fluorescence curves appeared to exhibit a small apparent difference in peak time (**Fig. 3B**). However, no difference in peak time was observed when calculating values across individual participants (**Fig. 3E**).

This effect can be explained by considering how the mean curves and individual parameters are calculated. Even a small number of high intensity curves with late peaks will shift the mean fluorescence vs. time curve (**Fig. 3B**) up and to the right, generating a later peak. However, the intensity of an individual curve does not affect the peak time extracted from it and, hence, intensity does not have an impact on the observed distribution of peak times (shown in **Fig. 3E**). This means that the average of the peak times extracted from each individual curve (**Fig. 3E**) and the peak time of the average curve (**Fig. 3B**) can have different values (as observed here). Importantly, however, the distribution (and average) of the peak times extracted from each individual curve (**Fig. 3E**) is representative of the true distribution of peak times in the dataset.

Hence, these results demonstrate that there was no difference in peak time between the UK and Zambian cohorts (as shown in **Fig. 3E**). This implies that there were no differences between the two groups in factors affecting the rate of uptake of fluorescein (e.g. gastric emptying, gut motility, etc.).

#### 4.2. Supplementary Note 2: Interpretation of peak AUC values

The parameters peak AUC, peak AUC / slope, and peak AUC * peak time exhibited no statistically significant differences between groups (**Fig. S6**). Interestingly, we have previously observed differences in these parameters between differing permeability states [3, 4], but this effect was not replicated here. This is likely due to the similar peak time values observed between groups in this study – i.e. peak time influences all of the parameters shown in **Fig. S6** and therefore acts to limit variability between groups in all cases.

#### 4.3. Supplementary Note 3: Demographic analysis

The data presented in **Fig. 4** demonstrates significant differences between the UK and Zambian cohorts in certain demographic groups. In particular, we observed significant differences between Group 1 (UK) and Group 2 (Zambia) for males (but not for females), for participants aged over 35 (but not under 35), and for participants in both BMI categories (>25 and <25) (**Fig. 4B-D**).

This potentially indicates that the differences in intestinal barrier function between UK and Zambian participants may be driven by differences in specific demographic groups. The data also suggests the possibility of differences in intestinal barrier function between different demographics. For example, although not statistically significant, **Fig. 4B&D** indicate lower peak intensity (i.e. lower intestinal permeability) in males than in females and higher peak intensity in BMI>25 than in BMI<25 in both groups. While this indicates potential demographic differences in gut function, it is important to note the small sample size used here. Particularly for Group 1, sample numbers are low when split according to demographics (e.g. only 4 female participants in Group 1; **Fig. 4B**), indicating that these observations should be interpreted with caution.
